# Heightened innate immunity may trigger chronic inflammation, fatigue and post-exertional malaise in ME/CFS

**DOI:** 10.1101/2025.07.23.25332049

**Authors:** Xiaoyu Che, Amit Ranjan, Cheng Guo, Keming Zhang, Rochelle Goldsmith, Susan Levine, Kegan J. Moneghetti, Yali Zhai, Liner Ge, Nischay Mishra, Mady Hornig, Lucinda Bateman, Nancy G. Klimas, Jose G. Montoya, Daniel L. Peterson, Sabra L. Klein, Oliver Fiehn, Anthony L. Komaroff, W. Ian Lipkin

## Abstract

Myalgic encephalomyelitis/chronic fatigue syndrome (ME/CFS) is characterized by unexplained fatigue, post-exertional malaise (PEM), and cognitive dysfunction. ME/CFS patients often report a prodrome consistent with infection. We present a multi-omics analysis based on plasma metabolomic and proteomic profiling, and immune responses to microbial stimulation, before and after exercise. We report evidence of an exaggerated innate immune response after exposures to microbial antigens; impaired energy production involving the citric acid cycle, beta-oxidation of fatty acids, and urea cycle energy production from amino acids; systemic inflammation linked with lipid abnormalities; disrupted extracellular matrix homeostasis with release of endogenous ligands that promote inflammation; reduced cell-cell adhesion and associated gut dysbiosis; complement activation; redox imbalance reflected by disturbances in copper-dependent antioxidant pathways and dysregulation of the tryptophan-serotonin-kynurenine pathways. Many of these underlying abnormalities worsened following exercise in ME/CFS patients, but not in healthy subjects; many abnormalities reinforced each other and several were correlated with the intensity of symptoms. Our findings may inform targeted therapeutic interventions for ME/CFS and PEM.

## Introduction

Myalgic encephalomyelitis/chronic fatigue syndrome (ME/CFS) is a debilitating disorder of at least six months’ duration characterized by unexplained fatigue, post-exertional malaise (PEM), cognitive impairment, and orthostatic intolerance. Many people report myalgias and gastrointestinal (GI) dysfunction^1^. Its global prevalence ranges between 0.4 and 2.5%. The United States alone has 2.5 million cases, and an annual economic burden of up to 51 billion dollars. ME/CFS is more common in females than males.^2^ The pathophysiology of PEM is unexplained; however, immune dysregulation and metabolic disturbances are described following exercise. Although no single agent is consistently implicated, patients often report a prodrome consistent with infection, including infection with betacoronaviruses implicated in SARS, MERS, and COVID-19 (the latter often called “Long-COVID”).^3^ The lack of convergence in efforts to identify etiological agents suggests that generic host responses to multiple microbial triggers may be important in its pathogenesis, and that ME/CFS and other post-infectious syndromes may represent a broader category.^4,5^ We report here a multi-omics analysis in patients with ME/CFS and healthy controls (HC), before and 24 hours after exercise, that suggests models for understanding the pathogenesis of chronic inflammation, fatigue, and PEM in ME/CFS.

## Results

### Study population and analytical datasets

Study subjects were 56 ME/CFS cases and 52 HC recruited in New York and California, with 47 case-control pairs matched for sex, age (±5 years), race/ethnicity, geography, date-of-exercise and blood sampling (±12 weeks), diagnosed using the CDC/Fukuda and Canadian consensus criteria.^1,6^ (**Table 1**, **Table S1, S2, Supplementary Materials 2.1, 2.2** for study population characteristics and analytical datasets).

**Table 1.** Subject characteristics. 1 Prior physician diagnosed irritable bowel syndrome, self reported on the questionnaire. 2 Multidimensional Fatigue Inventory; scored on 0−100 scale with 0 = no fatigue and 100 = greater fatigue. 3 SD: standard deviation 4 For categorical variables, p-values were derived from Chi-squared tests; For continuous variables, p-values were derived from from Wilcoxon rank-sum tests.

| Table 1. Subject characteristics. |  | Metabolomics & Truculture |  |
| --- | --- | --- | --- |
| Subject Characteristics |  | ME/CFS (n=56) | HC (n=52) |
| Sex | Female | 37 | 37 |
|  | Male | 19 | 15 |
| Age | Mean ± SD<sup>4</sup> | 46.1 ± 12.8 | 47.8 ± 13.7 |
|  | Median (Range: Min - Max) | 46.3 (21.0 - 70.0) | 50.5 (22.97 - 70.0) |
| Race and Ethnicity | White and not Hispanic | 47 | 41 |
|  | Hispanic | 4 | 5 |
|  | Not White and not Hispanic | 5 | 6 |
| Site of Collection | New York, NY | 36 | 34 |
|  | Palo Alto, CA | 20 | 18 |
| sr-IBS Comorbidity<sup>1</sup> | Yes | 10 | 1 |
|  | No | 46 | 51 |
| BMI | Mean ± SD<sup>5</sup> | 25.3 ± 4.9 | 23.8 ± 4.1 |
|  | Median (Range: Min - Max) | 24.6 (17.4 - 38.7) | 22.8 (17.2 - 33.1) |
| Duration of Illness | ≥ 3 years | 42 | n/a |
|  | < 3 years | 13 | n/a |
| MFI Scales<sup>2</sup><br>Mean Score ± SD<sup>3</sup><br>before exercise | General Fatigue | 69.4 ± 12.0 | 19.0 ± 14.6 |
| | Mental Fatigue | 74.0 ± 20.1 | 16.1 ± 14.3 |
| | Physical Fatigue | 84.2 ± 17.4 | 11.7 ± 11.6 |
| | Reduced Activity | 77.7 ± 23.1 | 12.1 ± 11.0 |
| | Reduced Motivation | 46.9 ± 25.0 | 13.1 ± 13.7 |
| MFI Scales<sup>2</sup><br>Mean Score ± SD<sup>3</sup><br>24 hours after exercise | General Fatigue | 69.6 ± 10.3 | 17.4 ± 15.5 |
| | Mental Fatigue | 74.3 ± 21.5 | 15.1 ± 16.2 |
| | Physical Fatigue | 84.8 ± 16.3 | 10.3 ± 11.7 |
| | Reduced Activity | 78.9 ± 22.2 | 12.3 ± 11.5 |
| | Reduced Motivation | 47.3 ± 25.7 | 10.8 ± 12.0 |
| MFI Scales<sup>2</sup><br>Mean Score ± SD<sup>3</sup><br>48 hours after exercise | General Fatigue | 90.8 ± 13.3 | 17.2 ± 14.8 |
| | Mental Fatigue | 73.2 ± 23.4 | 15.0 ± 16.6 |
| | Physical Fatigue | 86.3 ± 17.0 | 9.8 ± 10.5 |
| | Reduced Activity | 80.9 ± 22.6 | 11.8 ± 12.8 |
| | Reduced Motivation | 46.8 ± 18.3 | 35.8 ± 18.3 |

|  | Proteomics |  |  |
| --- | --- | --- | --- |
| *p*-value<sup>4</sup> | ME/CFS (n=36) | HC (n=34) | *p*-value<sup>4</sup> |
| 0.718 | 24 | 25 | 0.715 |
|  | 12 | 9 |  |
| 0.476 | 48.1 ± 13.1 | 49.9 ± 13.9 | 0.518 |
|  | 50.1 (21.0 - 70.0) | 51.0 (23.5 - 70.0) |  |
| 0.793 | 30 | 27 | 0.885 |
|  | 3 | 4 |  |
|  | 3 | 3 |  |
| 1.000 | 36 | 34 | n/a |
|  | 0 | 0 |  |
| 0.016 | 6 | 0 | 0.039 |
|  | 30 | 34 |  |
| 0.112 | 25.1 ± 5.6 | 23.8 ± 4.6 | 0.456 |
|  | 24.3 (17.4 - 38.7) | 22.6 (17.2 - 33.1) |  |
| n/a | 23 | n/a | n/a |
|  | 12 | n/a |  |
| <0.001 | 70.6 ± 13.3 | 20.1 ± 14.9 | <0.001 |
| <0.001 | 72.2 ± 20.2 | 15.6 ± 14.8 | <0.001 |
| <0.001 | 83.6 ± 15.6 | 12.5 ± 11.7 | <0.001 |
| <0.001 | 74.5 ± 23.9 | 12.3 ± 10.2 | <0.001 |
| <0.001 | 49.0 ± 23.5 | 11.9 ± 13.5 | <0.001 |
| <0.001 | 68.2 ± 11.4 | 17.9 ± 16.7 | <0.001 |
| <0.001 | 72.5 ± 23.0 | 15.2 ± 17.1 | <0.001 |
| <0.001 | 83.9 ± 15.6 | 11.0 ± 11.5 | <0.001 |
| <0.001 | 75.8 ± 22.7 | 13.4 ± 11.9 | <0.001 |
| <0.001 | 51.2 ± 24.7 | 10.5 ± 11.5 | <0.001 |
| <0.001 | 89.9 ± 13.8 | 17.6 ± 14.8 | <0.001 |
| <0.001 | 71.0 ± 24.5 | 14.2 ± 16.6 | <0.001 |
| <0.001 | 84.9 ± 17.2 | 10.4 ± 9.7 | <0.001 |
| <0.001 | 78.7 ± 23.5 | 13.1 ± 13.6 | <0.001 |
| <0.001 | 46.2 ± 18.3 | 32.6 ± 9.3 | <0.001 |

Statistical analyses yielded inferences on both between-group and within-group differences in levels of molecular analytes (**Methods – Statistical Analyses**). To facilitate interpretation, we used “higher/lower”, “elevated/reduced”, or “accumulated/depleted” to describe significant between-group differences and used “increased/decreased” to describe significant within-group changes.

### Heightened cytokine responses to SEB and HKCA in ME/CFS

We used the TruCulture *ex-vivo* system^7^ to ask whether expression of inflammatory mediators, before and after exercise, differed between ME/CFS and HC following exposure to superantigens and mimics of viral, bacterial and yeast infection: *Staphylococcus aureus* enterotoxin type B (SEB), heat-killed *Candida albicans* (HKCA), lipopolysaccharide (LPS) and polyinosinic:polycytidylic acid (poly I:C). In ME/CFS, levels of CXCL5, GM-CSF, IL-1β, IL-2, IL-6, IL-8, IL-23, and TNF-α were elevated before exercise in blood exposed to SEB (**Fig. 1A**, **Table S8E**). Similar elevations were seen with IFN-γ, IL-13, and IL-17 (*p*_adj_<0.10). SEB drives T-cell expansion by crosslinking T-cell receptors (TCRs) with major histocompatibility complex class II molecules (MHCIIs).^8^ ME/CFS cytokine profiles indicate elevations in superantigen-induced T-cell-mediated immune responses.

**Figure 1.**
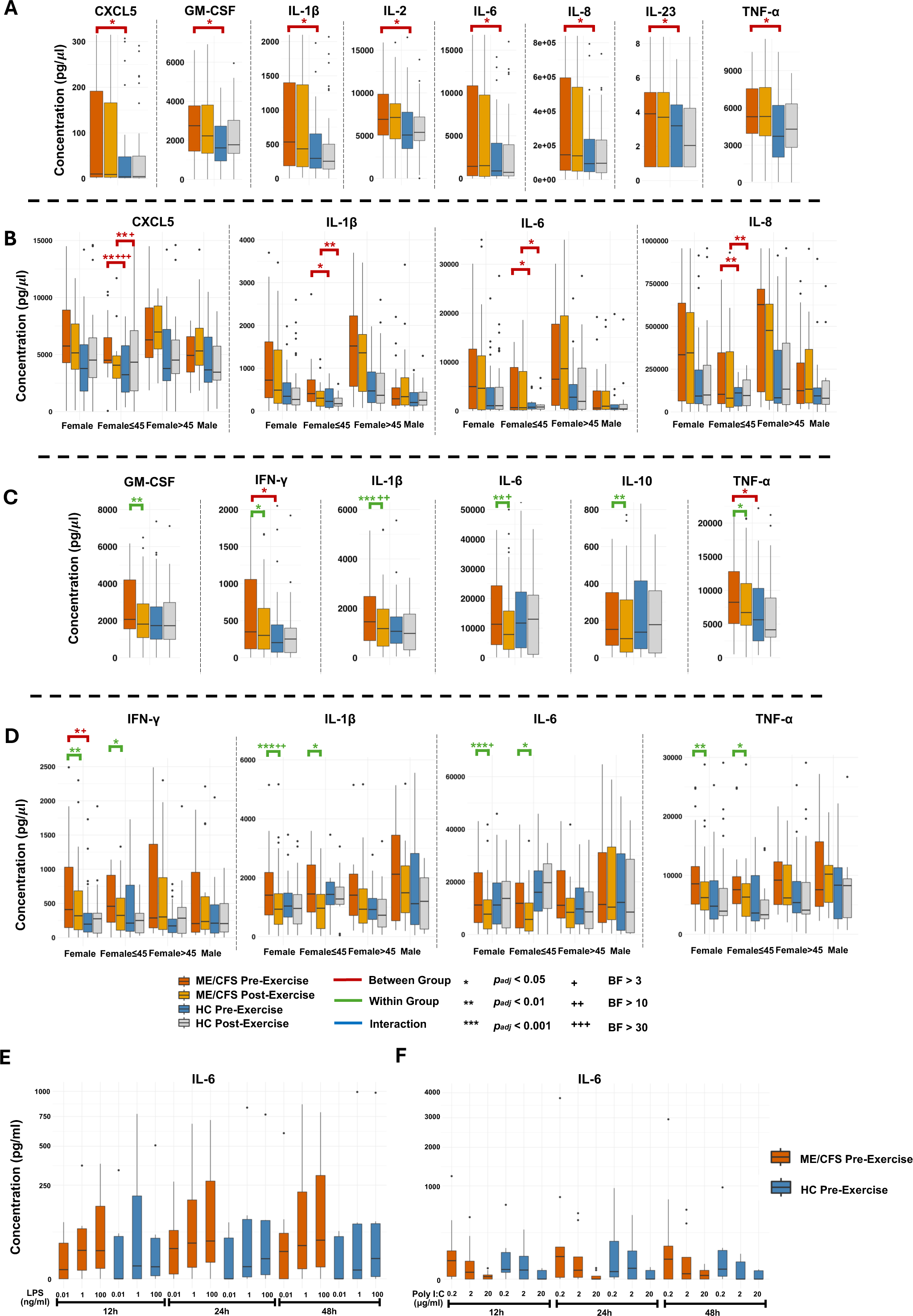
Inflammatory responses to microbial triggers. **(A). SEB ME/CFS vs HC**: Box plots showing concentrations of cytokines (pg/µl) in blood exposed to SEB before and 24 hours after exercise. Comparisons employed linear mixed-effect models (LMMs) paired with Bayesian analyses, adjusted for age, sex, race/ethnicity, geography, body mass index (BMI), and self-reported irritable bowel syndrome (sr-IBS). The full TruCulture cytokine results are reported in **Table S8**. **(B). SEB Sex- and age-stratified analysis:** Box plots showing sex-and age-stratified concentrations of cytokines (pg/µl) in blood exposed to SEB before and 24 hours after exercise. Comparisons employed linear mixed-effect models (LMMs) paired with Bayesian analysis in females and males, separately. The full TruCulture cytokine results are reported in **Table S8**. **(C). HKCA ME/CFS vs HC:** Box plots showing concentrations of cytokines (pg/µl) in blood exposed to HKCA before and 24 hours after exercise. Comparisons employed linear mixed-effect models (LMMs) paired with Bayesian analyses, adjusted for age, sex, race/ethnicity, geography, body mass index (BMI), and self-reported irritable bowel syndrome (sr-IBS). The full TruCulture cytokine results are reported in **Table S8**. **(D). HKCA Sex- and age-stratified analysis:** Box plots showing sex-and age-stratified concentrations of cytokines (pg/µl) in blood exposed to HKCA before and 24 hours after exercise. Comparisons employed linear mixed-effect models (LMMs) paired with Bayesian analysis in females and males, separately. The full TruCulture cytokine results are reported in **Table S8**. **(E). Dose- and time-dependent responses to LPS:** Box plots showing concentrations of IL-6 (pg/µl) induced in PBMCs before exercise in ME/CFS and HC after LPS stimulation. The doses were 0.01, 1 and 100 ng/ml of LPS and measured over 12, 24 and 48 hours post stimulation. **(F). Dose- and time-dependent responses to poly I:C:** Box plots showing concentrations of IL-6 (pg/µl) induced in PBMCs before exercise in ME/CFS and Controls after poly I:C stimulation. The doses were 0.2, 2 and 20 μg/ml of poly I:C and measured over 12, 24 and 48 hours post stimulation. SEB, staphylococcus aureus enterotoxin type B; HKCA, heat-killed Candida albicans; LPS, lipopolysaccharide; poly I:C, polyinosinic:polycytidylic acid.

Cytokine responses to SEB were greater in females (**Fig. 1B, Table S8E**), concordant with sex differences in both innate and adaptive immunological responses reported in microbial infections.^9^ We observed age-specific cytokine responses to SEB in female ME/CFS, that may reflect differences in levels of gonadal steroids, particularly the lower levels of 17β-estradiol (E2) seen in older women. We measured levels of E2 and testosterone in the pre-exercise plasma samples (**Table 2**). In response to SEB, female subjects with ME/CFS <45 years of age (YoA) had significantly higher levels of CXCL5, IL-1β, IL-6, and IL-8 than female HC <45 YoA before and after exercise; levels of IL-23 were elevated before exercise. Although female ME/CFS >45 YoA had higher levels of pro-inflammatory cytokines than older female HC, differences were not significant (**Fig. 1B**). Plasma levels of testosterone were negatively associated with age in females (**Table 2**). No significant association was observed between age and sex hormone levels in males after body mass index (BMI) adjustment.

**Table 2.** Sex-specific associations of plasma sex steroid levels with ME/CFS, age and BMI. Linear regression models were fitted with levels of each sex steroid as the dependent variable. The binary ME/CFS status, age and BMI were included as independent variables. Variables of age and BMI were standardized using z-scores. The interaction terms of ME/CFS x age were explored but were not included due to non-significance. Eight participants (7 women, 1 man) took hormone supplements or birth control measures (oral pills or intrauterine devices) and were excluded from this analysis. BMI body mass index.

|  |  | Female |  |  |  |
| --- | --- | --- | --- | --- | --- |
|  |  | Coefficient | 95% CI |  | p-value |
| Testosterone (pg/mL) | ME/CFS | 1.74 | -36.50 | 39.97 | 0.929 |
|  | Age | **-24.54** | **-43.72** | **-5.36** | **0.015** |
|  | BMI | 6.90 | -12.68 | 26.47 | 0.492 |
| Estradiol (pg/mL) | ME/CFS | 2.35 | -26.50 | 31.21 | 0.874 |
|  | Age | **-22.49** | **-36.97** | **-8.01** | **0.003** |
|  | BMI | -10.15 | -24.93 | 4.62 | 0.183 |

| Male |  |  |  |
| --- | --- | --- | --- |
| Coefficient | 95% CI |  | p-value |
| 175.88 | -783.67 | 1135.42 | 0.722 |
| 45.52 | -457.38 | 548.41 | 0.860 |
| **-640.97** | **-1146.08** | **-135.86** | **0.019** |
| -2.77 | -6.75 | 1.20 | 0.182 |
| 1.84 | -0.24 | 3.93 | 0.093 |
| 1.07 | -1.03 | 3.16 | 0.326 |

In response to HKCA, ME/CFS subjects had elevated levels of IFN-γ and TNF-α before exercise compared to controls. Baseline levels of IL-1β were also elevated but did not reach significance (*p*_adj_=0.073). Host defense against *C. albicans* commences with the activation of NF-κB, AP-1 (JUN and FOS) and MAPK signaling cascades, with concomitant release of IFN-γ and TNF-α.^10–12^ The pre-exercise cytokine elevations in response to HKCA were similar in females and males with ME/CFS (**Fig. 1D**). After exercise, levels of GM-CSF, IFN-γ, IL-1β, IL-6, IL-10, and TNF-α in response to HKCA were decreased in females with ME/CFS, but not in males (**Fig. 1C, Table S8C**). The reductions were similar between younger and older females with ME/CFS (**Fig. 1D**). We speculate that decreased cytokine levels in response to HKCA, as well as the lack of exercise-induced changes in cytokine levels in response to SEB, may be the result of immune exhaustion or dysregulation in subgroups of immune cells.^13^

These findings suggest hypersensitivity of ME/CFS to microbial stimulus of innate and adaptive immunity. The observations also led us to examine the differences in plasma metabolomics and proteomics between the two age-subgroups within female ME/CFS. We discuss findings on differences between ME/CFS and HC here and findings within subgroups of ME/CFS stratified by sex, age, and duration of illness^14^ (**Supplementary Materials 1.2** for subgroup stratification) in **Supplementary Materials 2.3 (Fig. S2-S7)**.

### Sensitivity of cytokine responses to LPS and poly I:C

TruCulture preparations exposed to LPS and poly I:C revealed higher secretion of pro-inflammatory cytokines compared to no stimulation, in both ME/CFS and HC (**Table S1C**). However, we did not observe case-control differences (**Fig. S1**, **Table S8B & S8D**). The TruCulture system employs a single dose of stimulant and measures cytokine expression at a single time point. We speculated that differences in sensitivity to LPS and polyI:C between ME/CFS and HC might be obscured by saturation of their cognate Toll-like receptors. Our challenge in formally testing this hypothesis was the limited number of remaining PBMC samples from ME/CFS and HC after earlier experiments. As a result, we were unable to assess post-thaw viability; however, PBMC aliquots from ME/CFS and HC were collected and cryopreserved under identical conditions and were only thawed immediately prior to use. We recovered and tested pre-exercise PBMCs from ME/CFS (n=8) and controls (n=7) with three doses of LPS (0.01, 1 and 100 ng/ml) and poly I:C (0.2, 2, 20 μg/ml) for 12, 24 and 48 hours and measured induction of IL-6. We observed dose- and time-dependent IL-6 responses to both LPS and poly I:C stimulation at 12, 24 and 48 hours of incubation (**Fig. 1E & 1F**).

### Proteomic correlates of altered activation of innate immunity

At baseline, plasma levels of colony stimulating factor 1 (CSF1) and low-density lipoprotein receptor-related protein 8 (LRP8) were lower in ME/CFS (**Fig. 2E**), consistent with earlier reports.^14^ CSF1 activates dendritic cells (DCs), monocytes, and macrophages by modulating cell surface receptors.^15^ Lower levels of CSF1 may indicate inadequate activation and maturation of immune cells, impairing resolution of inflammation. Levels of protein C (proC) and basic leucine zipper transcriptional factor ATF-like 3 (BATF3) were elevated in ME/CFS both before and after exercise (**Fig. 2E, 2F**). proC is a pleiotropic circulating protein that primarily functions as an anticoagulant but also contributes to endothelial and epithelial barrier integrity by participating in vascular basement membrane formation.^16,17^ proC also activates the platelets via interacting with apolipoproteins (LRP8).^16^ BATF3 is essential for the development of CD8a+ and CD103+ dendritic cells.^18^ Post-exercise levels of endogenous retrovirus group V member 1 (ERVV1) were elevated in ME/CFS. Endogenous retroviruses activation has been associated with inflammation and neurological disorders.^19^

**Figure 2.**
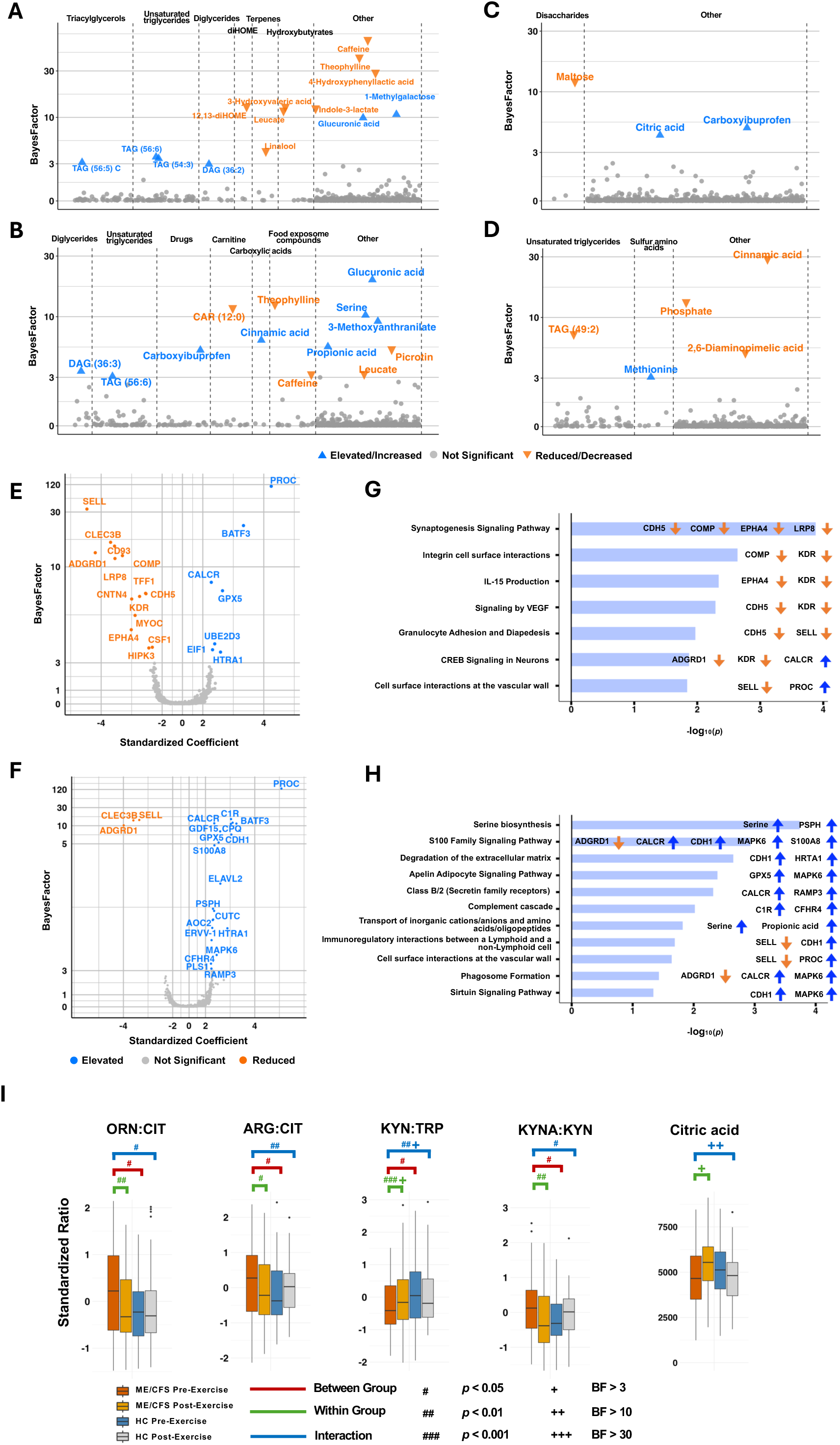
Multi-omics analyses comparing ME/CFS vs. HC. **(A and B). Metabolomics**. BayesFactors (BFs) of 913 metabolites in plasma comparing ME/CFS vs. HC (**A**) before and (**B**) 24 hours after exercise, using linear mixed-effect models (LMMs) paired with Bayesian analyses. The LMM models were adjusted for age, sex, race/ethnicity, geography, body mass index (BMI), and self-reported irritable bowel syndrome (sr-IBS). Significant metabolites were labeled (significance defined as 1. FDR adjusted *p*<0.05, or 2. BF>3 and 95% HDI does not cover 0, or 3. BF_combined_>10 and HDI_combined_ does not cover 0). If BF_combined_>10 and BF_combined_>BF, BF_combined_ is plotted, otherwise, BF is plotted. Gray dots at the bottom of the figure represent individual metabolites that were not significantly altered between ME/CFS and HC. The significant chemical clusters identified by ChemRICH were individually labeled. The significant metabolites in other nonsignificant clusters were labeled under “Other” on the right. The full metabolomic results, comparing ME/CFS and HC, are reported in **Table S3A**. **(C and D).** Scatter plots showing BayesFactors (BFs) associated with changes in plasma levels of metabolites from before to 24 hours after exercise in (**C**) ME/CFS and in (**D**) HC. The full metabolomic results, comparing ME/CFS and HC, are reported in **Table S3A**. **(E and F). Proteomics.** Volcano plots showing BayesFactors (BFs) of proteomic analytes in plasma comparing ME/CFS vs. HC (**E**) before and (**F**) after exercise, using linear mixed-effect models (LMMs) paired with Bayesian analyses. The LMM models were adjusted for age, sex, race/ethnicity, geography, body mass index (BMI), and self-reported irritable bowel syndrome (sr-IBS). Significant proteins were labeled (significance defined as 1. FDR adjusted *p*<0.05, or 2. BF>3 and 95% HDI does not cover 0). Gray dots at the bottom of the figure represent individual proteins that were not significantly different between ME/CFS and HC. The full proteomic results, comparing ME/CFS and HC, are reported in **Table S6A**. **(G and H). Integrative enrichment.** Ingenuity pathway analyses (IPA), combining plasma metabolomic and proteomic results, comparing ME/CFS vs. HC (**G**) before and (**H**) after exercise. The full IPA results, comparing ME/CFS and HC, are reported in **Table S7A**. **(I). Citric acid and ratios of metabolites.** Box plots showing levels of citric acid and standardized ratios of ornithine (ORN) to citrulline (CIT), arginine (ARG) to citrulline (CIT), kynurenine (KYN) to tryptophan (TRP) and kynurenic acid (KYNA) to kynurenine (KYN) in ME/CFS and HC before and 24 hours after exercise. Comparisons employed linear mixed-effect models (LMMs) paired with Bayesian analyses, adjusted for age, sex, race/ethnicity, geography, body mass index (BMI), and self-reported irritable bowel syndrome (sr-IBS). The full results from ratio analysis, comparing ME/CFS and HC, are reported in **Table S4A**.

### Abnormalities in cell-extracellular matrix proteins and cell-cell interactions

Integrative Ingenuity Pathway Analysis (IPA) revealed that levels of analytes in pathways associated with integrin cell surface interactions, VEGF signaling, and leukocyte adhesion and diapedesis were reduced before exercise; levels of analytes in the extracellular matrix protein (ECM) degradation pathway were elevated after exercise (**Fig. 2G, 2H, Table S7A**). Baseline levels of CD93 and cartilage oligomeric matrix protein (COMP) were lower; baseline levels of high-temperature requirement serine protease A1 (HTRA1) were higher; pre- and post-exercise levels of tetranectin (CLEC3B) were lower (**Fig. 2E, 2F**). HTRA1 degrades ECM proteins, including fibronectin, decorin, type II collagen, elastin, and aggrecan.^20^ CD93 is a c-type lectin that modulates immune cell migration to the endothelial cell surface and has anti-inflammatory properties.^21^ Tetranectin, another c-type lectin, regulates ECM remodeling through binding to plasminogen or high mobility group box 1 (HMGB1).^22^ ECM protein COMP, produced by cells of connective tissue and platelets, protects against age-related vascular dysfunction and maintains vascular homeostasis through its secretion and organized incorporation into the ECM.^23,24^

In ME/CFS, levels of L-selectin (SELL) and adhesion G-protein coupled receptor (GPCR) D1 (ADGRD1) were reduced both before and after exercise; baseline levels of cadherin-5 (CDH5/VE-cadherin) and kinase insert domain receptor (KDR) were reduced (**Fig. 2E, 2F**). SELL is a leukocyte cell surface molecule that plays roles in immune surveillance through leukocyte adhesion to endothelial cells and facilitating homing of activated T cells.^25^ Glycoprotein CDH5 is localized on the junctions of endothelial cells and regulates inflammatory responses at adherens junctions.^26^ The gene encoding ADGRD1, GPR133, couples to Gαs and increases intracellular levels of cAMP and is thereby involved in neuronal activity and cognitive functions.^27^ KDR is the primary receptor for VEGF.^28^ KDR and VEGF expressed on endothelial cells have pro-inflammatory properties and mediate T-cell trafficking.^28^ KDR and VEGF expression can also be induced in T-cells by hypoxia, and the VEGF-KDR complex promotes Th1 differentiation. Lower plasma levels of SELL, CDH5, ADGRD1, and KDR suggest impaired immune surveillance and dysregulated immune activation.

### Disrupted GI mucosal integrity

2,6-Diaminopimelic acid (DAP) is an important component of peptidoglycans. After exercise, plasma levels of DAP were decreased in HC, but not in ME/CFS (**Fig. 2D**). Plasma levels of leucate (2-Hydroxy-4-methylpentanoic acid), another microbial metabolite, were reduced in ME/CFS before and after exercise (**Fig. 2A, 2B**). Furthermore, baseline levels of citrulline (CIT) trended toward reduction in ME/CFS (**Table S3A, see urea cycle below**). CIT, primarily produced by enterocytes in the gut, is considered a biomarker for functional enterocyte mass and intestinal mucosal integrity.^29^ Baseline levels of TFF1, secreted by mucus producing cells and important for rapid healing of barrier through restitution, were reduced in ME/CFS (**Fig. 2E**).^30^ These observations indicate compromised GI-barrier consistent with dysbiosis in ME/CFS.

### Dysregulated calcium signaling and complement activation after exercise

After exercise, analytes in the S100 family and Class B/2 signaling pathways were elevated in ME/CFS (**Fig. 2H**). Levels of calcitonin receptor (CALCR) were higher in ME/CFS both before and after exercise; levels of receptor activity-modifying protein 3 (RAMP3) and S100A8 were higher after exercise (**Fig. 2E, 2F**). CALCR (GPCR) binds calcitonin to regulate calcium homeostasis. Its binding preference is modulated by RAMPs. S100A8 is a small calcium-binding protein expressed in leukocytes and neutrophils that accelerates inflammation during infection or in immune hypersensitivity diseases.^31^ These findings reflect abnormalities in calcium signaling after exercise that may contribute to inflammation and neuronal dysfunction in PEM. We also observed complement activation in ME/CFS after exercise (**Fig. 2H**). Levels of C1R and complement factor H-related protein 4 (CFHR4) were elevated in ME/CFS after exercise (**Fig. 2F**). C1R is one of the initial components activating complement reactions through the classical pathway; CFHR4 is a regulatory protein required to form active C3 convertase, via interaction with C3b, in the alternative pathway.^32^ Prolonged complement activation can lead to fatigue, systemic inflammation, and cognitive impairment, consistent with PEM.

### Mitochondrial dysfunction

Mitochondrial dysfunction has been well-documented in ME/CFS at rest, with reported impairments in electron transport chain (ETC) activity and reduced ATP synthesis^33–35^, although not all studies have reported these findings^36^. In ME/CFS subjects reported here, we found higher plasma levels of growth differentiation factor 15 (GDF15) after exercise (**Fig. 2F**).

Activation of the integrated stress response pathway induces the secretion of GDF15 which serves as a biomarker of mitochondrial dysfunction.^37–39^ Citric acid (citrate) is an important intermediate in the tricarboxylic acid (TCA) cycle. Although Yamano et al. reported lower plasma levels of citrate in ME/CFS at rest^40^, there are no published data on citrate levels after exercise. Here, plasma levels of citrate were increased after exercise in ME/CFS but decreased in HC (**Fig. 2C, 2I**). The citrate pool is maintained through metabolic production of citrate via the TCA cycle, dietary intake, renal reabsorption, and bone resorption.^41^ Given the fasting state during sampling and the within-subject comparisons, we speculate that the post-exercise increase in plasma citrate levels observed in ME/CFS may reflect reduced mitochondrial flux and altered metabolism. After exercise, levels of phosphate were decreased in HC, but not in ME/CFS (**Fig. 2D**). If we interpret this finding as indicating phosphate utilization for ATP generation via the TCA cycle and downstream pathways in HC, the absence of a comparable decrease in ME/CFS is consistent with mitochondrial dysfunction.

We found evidence of baseline mitochondrial dysfunction primarily in subgroups of ME/CFS (**Supplementary Materials 2.3.8** for subgroup-specific effects in mitochondrial dysfunction and lipid abnormalities), including impairments in mitochondrial respiratory chain and oxidative phosphorylation (OXPHOS) that are consistent with previous reports.^33–35^

### Abnormalities in lipid metabolism

We and others have reported elevated baseline levels of triglycerides in ME/CFS.^42,43^ Here, ChemRICH analysis revealed higher plasma levels of triglycerides and diglycerides before and after exercise (**Fig. 2A, 2B, Table S5**). ChemRICH analysis also revealed lower levels of carnitines, particularly acylcarnitine CAR (12:0), after exercise (**Fig. 2B, Table S5**). Carnitines are essential in the transport of long-chain FAs from cytoplasm to mitochondria for β-oxidation. Acylcarnitines are exported from mitochondria during nutritional overload (or reduced β-oxidation capacity).^44^ We previously reported reduced baseline plasma levels of acylcarnitines in ME/CFS in two independent cohorts.^43^ Acylcarnitine depletions before and after exercise indicate a lower flux of mitochondrial FA for β-oxidation, further contributing to triglyceride accumulation.

Before exercise, ME/CFS had lower plasma levels of diHOMEs, particularly 12,13-diHOME (**Fig. 2A**). 12,13-diHOME, a cold-induced lipokine, promotes the translocation of transporters FATP1 and CD36 to the plasma membrane of brown adipocytes, thereby enhancing FA uptake.^45^ In healthy subjects, levels of 12,13-diHOME increase after acute exercise and return to basal levels after 1 hour.^45^ In ME/CFS, levels of both 12,13-diHOME and its precursor linoleic acid (LA) were increased after exercise compared to HC. The shift is borderline non-significant for 12,13-diHOME but significant for LA (**Table S3A**).

### Abnormalities in tryptophan and neuronal metabolism

Tryptophan (TRP) is an essential amino acid critical for protein synthesis that is metabolized through the serotonin and kynurenine (KYN) pathways. The neurotransmitter serotonin regulates mood, appetite, sleep, and cognition. Reductions in circulating serum levels of serotonin are reported in ME/CFS. We found that, irrespective of antidepressant medication, KYN:TRP ratios were lower in ME/CFS before exercise and increased after exercise (**Fig. 2I, Table S4**). Increased KYN:TRP ratios suggest overactivation of KYN pathway in ME/CFS after exercise that could divert TRP from the serotonin pathway, leading to serotonin deficiency and overproduction of KYN pathway metabolites. The ratios of kynurenic acid (KYNA) to KYN, an indirect index for kynurenine aminotransferase (KAT) activity, were higher in ME/CFS before exercise and decreased after exercise (**Fig. 2I**). Simonato et al. measured higher KYNA:KYN ratios in serum of ME/CFS patients at rest.^46^ Whereas KYN exhibits depressogenic properties, KYNA is neuroprotective.^47^ Post-exercise KYNA reduction relative to KYN indicates reduced KAT activity and elevated levels of 3-methoxyanthranilate (3-hydroxy anthranilate derivative) (**Fig. 2B**) lead to neurotoxicity and neuroinflammation.^48^

TRP can also be metabolized via the gut microbiota-derived indole pathway.^49^ The metabolomic analysis confirmed findings of depleted baseline levels of indole-3-lactate/indole lactic acid (ILA) in plasma of ME/CFS (**Fig. 2A**). ILA is derived from commensal bacterium, such as *Bifidobacterium*, and plays important roles in regulating inflammation.^49^ Decreased fecal levels of short chain fatty acid (SCFA) butyrate are reported in ME/CFS.^50^ We did not detect butyrate in plasma from ME/CFS or HC subjects; however, plasma levels of propionic acid (PPA), another SCFA, were elevated after exercise (**Fig. 2B, 4F**). PPA can disrupt mitochondrial biogenesis and induce neuroinflammation.^51^ It is also associated with cognitive decline during aging, suggesting that elevated PPA levels in ME/CFS may contribute to post-exercise impairments in cognitive function.^52,53^

Plasma proteomic findings also revealed abnormalities in proteins predominantly produced by neuronal cells. Before exercise, levels of Ephrin receptor A4 (EPHA4) and contactin 4 (CNTN4) were lower in ME/CFS (**Fig. 2E**). EPHA4 is highly expressed in brain regions with high synaptic plasticity and is implicated in learning and memory.^54^ CNTN4 is expressed in both peripheral and central nervous system and is involved in modulating signal transduction at synapses.^55^ After exercise, levels of ELAV-like RNA binding protein 2 (ELAVL2) were elevated (**Fig. 2F**). ELAVL2 regulates the post-transcriptional expression of key genes involved in neuronal development and function.^56^ In response to exercise, levels of Neurexin1 (NRXN1) were decreased in ME/CFS (**Table S6A**), indicating lower synaptic adhesion and neuronal transmission that may contribute to PEM-associated cognitive impairment.

### Urea cycle disruption

The activity of urea cycle is reflected in the ratios between pairs of intermediates ornithine (ORN), CIT, and arginine (ARG). Yamano et al. reported higher ORN:CIT ratios at rest in plasma of ME/CFS subjects.^40^ Before exercise, plasma ORN:CIT ratios were elevated in ME/CFS. ARG:CIT ratios were also higher (**Fig. 2I, Table S4**). Baseline accumulations of ARG and ORN relative to CIT may reflect impairments in the initial steps of urea cycle, where CIT is formed from ORN and carbamoyl phosphate.^40^ Following exercise, the ORN:CIT and ARG:CIT ratios decreased (**Fig. 2I).** These findings are consistent with impairments in the later stages of urea cycle, where CIT reacts with aspartic acid to form argininosuccinic acid, which is subsequently converted to ARG. Changes in the ratios between urea cycle intermediates were primarily driven by alterations in CIT levels (**see GI integrity**), possibly due to reduced activity of either CIT transporters or enzymes of urea cycle.

### Dysregulated xenobiotic metabolism

Glucuronidation in the liver can contribute to detoxification by removing toxins, xenobiotics, and endogenous compounds. This process may be confounded by gut β-glucuronidases produced by *Bacteroidetes* that cleave glucuronide conjugates, releasing free xenobiotic compounds and glucuronic acid that are utilized as energy sources. Fecal levels of *Bacteroidetes* are reported to be elevated in ME/CFS.^57^ Concordant with increased β-glucuronidase activity, we observed higher levels of glucuronic acid both before and after exercise (**Fig. 2A, 2B**). We used fasting plasma samples; thus, glucuronic acid accumulation is more likely to reflect dysbiosis and dysregulated xenobiotic metabolism than differences in diet.

### Proteomic correlates of cellular stress

Before exercise, levels of eukaryotic translation initiation factor 1 (EIF1) and ubiquitin-conjugating enzyme E2 D3 (UBE2D3) were higher in ME/CFS (**Fig. 2E**). EIF1 facilitates translational initiation during protein synthesis through interactions with the 40S ribosomal subunit. Elevated plasma levels may result from cellular leakage of EIF1 into circulation, indicating errors in translational initiation. The ubiquitin-proteasome system (UPS) maintains protein quality control and cellular homeostasis by tagging damaged and misfolded proteins for degradation. These findings are consistent with a hypometabolic state in ME/CFS at rest proposed by Naviaux et al.^58^

After exercise, levels of retina-specific copper amine oxidase (AOC2) and copper homeostasis protein cutC homolog (CUTC) were higher in ME/CFS (**Fig. 2F**). AOC2 is a copper-dependent enzyme that catalyzes oxidation of primary amines, including neurotransmitters such as dopamine and serotonin, into aldehydes, H_2_O_2_, and ammonia. CUTC facilitates intracellular copper transport and regulates copper homeostasis. Copper is a cofactor for the oxidant defense system, which includes superoxide dismutase (SOD), catalase (CAT) and glutathione (**Supplementary Materials 2.3.10, 2.3.11** for peroxisomal dysfunction and amino acid abnormalities). Elevated post-exercise levels of AOC2 and CUTC indicate increased oxidative stress due to the elevated generation of reactive oxygen species (ROS).^59^ Post-exercise levels of MAPK6 were also elevated (**Fig. 2F**). MAPK6, an atypical member of MAPK subfamily, is induced upon oxidative stress, leading to the production of IL-8 and the secretion of epithelial-derived factors in chemotaxis.^60^ In concert, these findings suggest a prolonged oxidative state in ME/CFS and an increase in oxidative stress levels following exercise.

### Plasma proteomic and metabolic abnormalities correlate with ME/CFS symptoms

Baseline levels of 12,13-diHOME were lower in ME/CFS than in HC. In both ME/CFS and HC, lower baseline levels of 12,13-diHOME correlated with higher Multidimensional Fatigue Inventory (MFI) scores of physical fatigue and reduced activity.^61^ Post-exercise levels of GDF15 were higher in ME/CFS than in HC. Higher post-exercise levels of GDF15 positively correlated with MFI scores of general fatigue, physical fatigue, and reduced activity (**Fig. 3A-3D**). Baseline levels of CNTN4, a molecule important in neuroplasticity, were reduced in ME/CFS. Reduced levels of CNTN4 correlated with higher general fatigue scores in ME/CFS. Reduced pre- and post-exercise levels of tetranectin (CLEC3B) in ME/CFS correlated with higher MFI scores of physical fatigue and reduced activity/motivation, respectively. Elevated post-exercise levels of S100A8 and C1r in ME/CFS correlated with MFI scores of reduced activity/motivation (**Fig. 3A, 3B**). These findings are consistent with dysregulation of ECM remodeling, calcium signaling, and complement pathways. Prioritizing interventions to modulate metabolic regulators and anti-inflammatory agents may mitigate multifactorial burden of ME/CFS. Inter-analyte correlations revealed distinct patterns between ME/CFS and HC (**Fig. 3E, 3F, Supplementary Materials 2.4** for analyte-analyte correlations).

**Figure 3.**
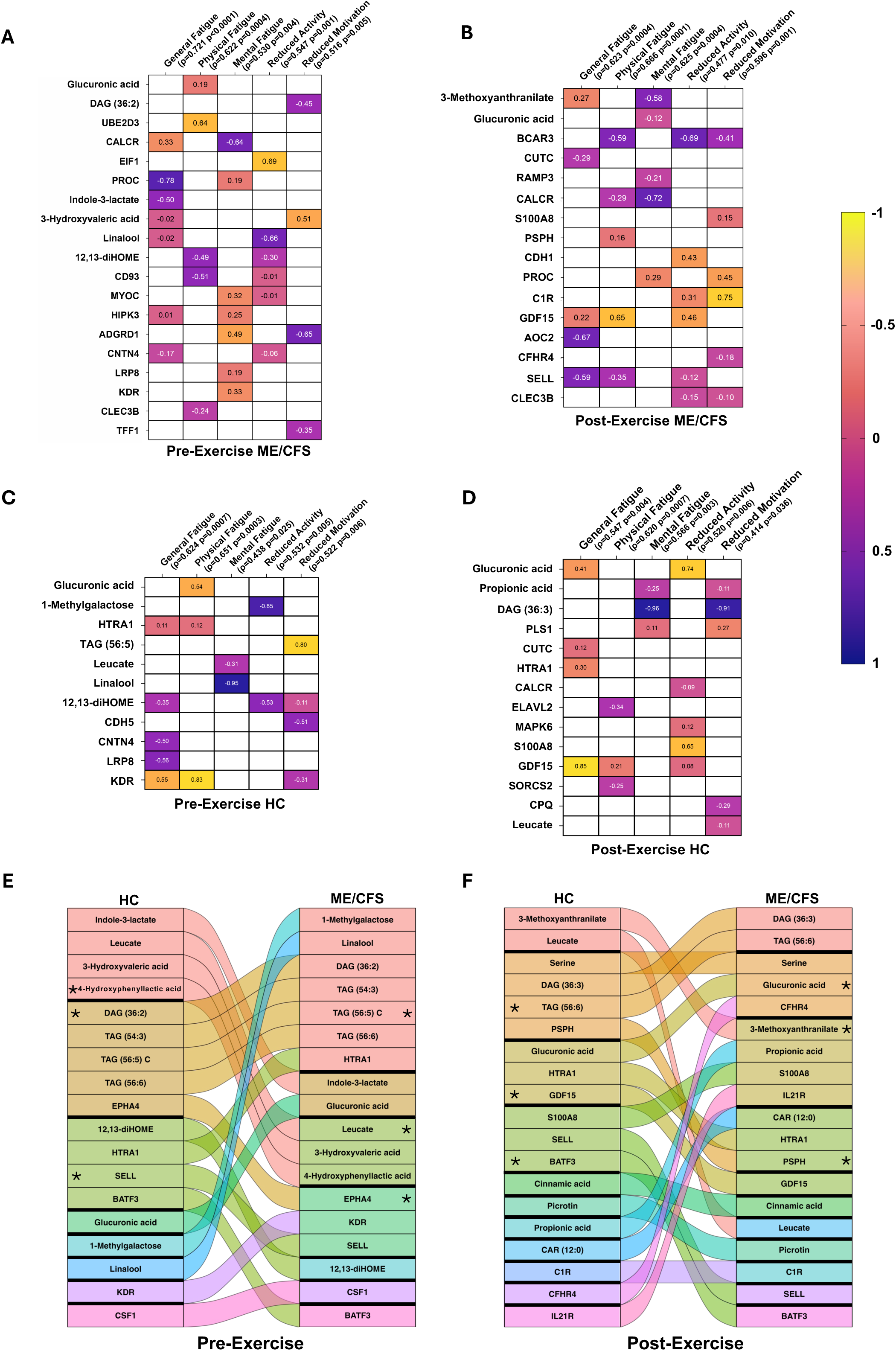
Correlations between metabolites, proteomic analytes and clinical fatigue scores. **(A -D) Analyte-Symptom correlations**. Heat maps showing correlations between significantly altered analytes and MFI symptom severity in ME/CFS (**A**) before and (**B**) after exercise, and in HC (**C**) before and (**D**) after exercise. Regularized canonical correlation analysis (RCCA) with Lasso penalty was used. Drug metabolites were excluded in this analysis. The full analyte-symptom correlation results are reported in **Table S9.** (**E and F**) **Analyte-analyte correlations**. Alluvial plots showing the spearman correlations, in ME/CFS and HC, separately, between plasma levels of metabolomic and proteomic analytes, which showed significant difference between ME/CFS and HC (**E**) before and (**F**) 24 hours after exercise. Drug metabolites were excluded in this analysis. Clustering was performed using the Louvain Method and the module centers (or hubs) were identified based on eigenvector centrality scores.

## Discussion

The sine qua non of ME/CFS are chronic fatigue and PEM. Metabolomic and proteomic analyses of plasma, before and 24 hours after exercise, revealed mechanistic insights into these symptoms. Although correlations between biomarkers for inflammation and disease severity have not been conclusively demonstrated, we and others have proposed that inflammation may be implicated in the pathogenesis of ME/CFS.^14,62–68^ Analyses of plasma and supernatant of whole blood exposed to innate immune stimuli confirmed immunological and metabolic abnormalities that can trigger inflammation. Experiments with PBMCs exposed to LPS and poly I:C revealed higher levels of IL-6 in ME/CFS than in HC. We found new metabolomic and proteomic markers of systemic inflammation referent to triglyceride accumulation, disrupted ECM homeostasis, and reduced cell-cell adhesion.

Although the majority of ME/CFS subjects report a prodrome consistent with infection, no single infectious agent has been implicated.^69^ This and reports of an ME/CFS-like syndrome in some patients with Long-COVID prompted us to consider a model for the pathogenesis of ME/CFS focused on host responses to microbial stimuli. In the TruCulture *ex vivo* system, immune stimulation with SEB and HKCA induced higher levels of pro-inflammatory cytokines in ME/CFS than in HC with sex- and age-specific effects. Although some findings did not reach statistical significance (*p*_adj_<0.10), the pattern of the data was consistent across pro-inflammatory cytokines (**Fig. 1A, 1B**). We posit that these differences were influenced by E2, a master regulator of inflammation inhibiting the release of pro-inflammatory cytokines, including IL-1β, IL-6, and IL-23. E2 also promotes expansion of Treg cells, and inhibits differentiation of Th17 cells and production of Th17 cytokines^9^. However, no case-control differences were found in cytokine responses to LPS and poly I:C. We speculated that the lack of differences reflected saturation of TLRs with the stimulant concentration and incubation time employed in the assays. Accordingly, we tested IL-6 responses in PBMCs exposed to various doses of LPS or poly I:C for 12, 24 or 48 hours. These experiments revealed differences in innate immune responses that were most evident at lower stimulant concentrations and at 24 hours of incubation. We found no differences between ME/CFS and HC in levels of TLR-3 or TLR-4 (**Table S6A**) or their correlate transcripts in analyses of bulk PBMC preparations (unpublished). This does not exclude the possibility of differences in subpopulations of PBMCs that might account for differences in sensitivity to superantigens or TLR agonists.

Mitochondria are important not only for energy production, but also for the regulation of inflammation. Mitochondrial dysfunction may induce triglyceride accumulation by reducing peroxisomal activity (**Supplementary Materials 2.3.10** for peroxisomal dysfunction), leading to lower carnitine shuttle function in muscle and impaired FA β-oxidation.^70^ Conversely, peroxisomal dysfunction has recently been shown to induce mitochondrial dysfunction, through a novel mechanism.^71^ We have previously demonstrated peroxisomal dysfunction in subjects with ME/CFS^40^. The bidirectional connection between peroxisomal dysfunction and mitochondrial dysfunction can become a vicious cycle. Such vicious cycles may contribute to the persistence of the symptoms of the illness.^72^

Accumulated triglycerides and diglycerides can, in turn, trigger inflammation, cellular stress, insulin resistance, cell-ECM disruption, and further mitochondrial dysfunction.^73^ Reduced levels of tetranectin, CD93, and COMP, along with elevated levels of HTRA1, suggest enhanced ECM degradation and disruption of ECM homeostasis resulting into release of endogenous ligands that promote inflammation(**Fig. 4A, 4B**).^74^ Reduced cell-cell adhesion, particularly in intestinal epithelium, may contribute to translocation of microbial products from the gastrointestinal tract to the systemic circulation that trigger innate immune responses and impair the function of tissue-resident innate lymphoid cells important for immune regulation.^75^ We and others have reported alterations in the gut microbiome and evidence of translocation of bacterial components from gut to circulation in ME/CFS.^50^ Butyrate, an important SCFA in maintaining GI mucosal integrity, and butyrate-producing bacteria are lower in the fecal samples of ME/CFS subjects.^50^ In this study, we found altered plasma levels of microbial metabolites—leucate, DAP, and PPA—that are associated with gut dysbiosis and may contribute to systemic inflammation.^51,76^ Reduced pre- and post-exercise levels of leucate in ME/CFS are compatible with our previous report that fecal levels of *R. torques* correlated with MFI symptom scores^50^. Consistent with PEM, we found that inflammation was exacerbated following exercise, as indicated by complement activation and altered calcium signaling. Prolonged complement activation may result in activation of immune cells, increased vascular permeability, and tissue damage. Functional impairments in platelets, monocytes, macrophages, DCs, NK cells, B-cells, and T-cells are reported in ME/CFS.^77^ Our findings with CSF, LRP8, BATF3 and proC further strengthened evidence for the role of innate immunity in ME/CFS pathogenesis.

**Figure 4.**
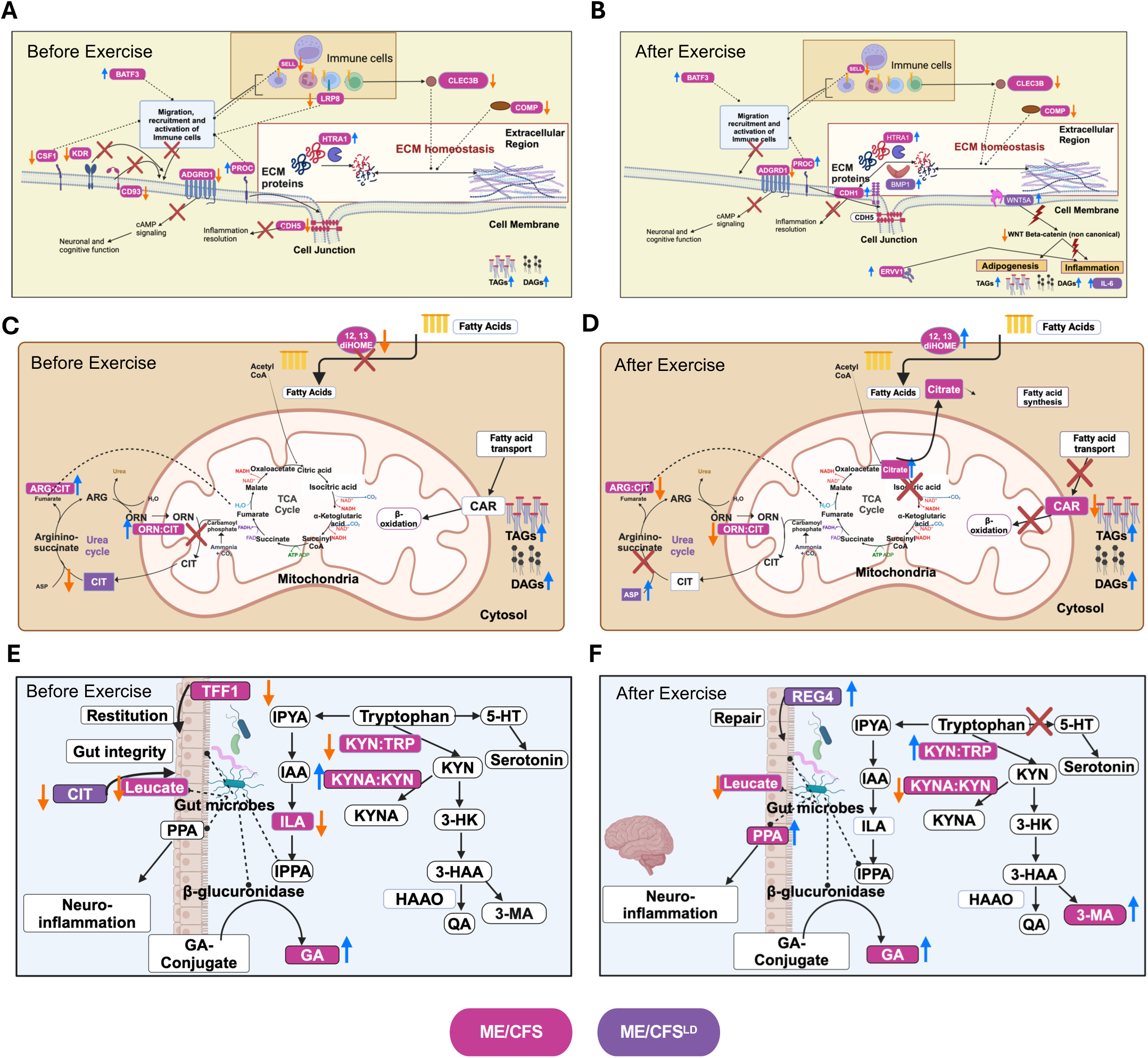
Potential mechanisms for inflammation, fatigue, and PEM. **(A and B). Mechanistic model of Inflammation (A) before and (B) after exercise. A.** Before exercise, plasma levels of TAGs, DAGs, and HTRA1 were elevated in ME/CFS; levels of SELL, CDH5, CD93, ADGRD1, CLEC3B, and COMP were lower. Elevated levels of TAGs and DAGs reflect inflammation. Reduced levels of SELL, CD93, ADGRD1, CSF1, KDR and CDH5 result in impaired recruitment of immune cells to tissues. Reduced levels of CLEC3B and COMP, as well as elevated levels of HTRA1, may disrupt ECM homeostasis. **B.** After exercise, plasma levels of TAGs, DAGs, HTRA1, and CDH1 were elevated in ME/CFS; levels of SELL, ADGRD1, CLEC3B, and COMP were reduced. Levels of IL-6, BMP1, and WNT5A were elevated in long duration (>3 years) ME/CFS. Elevated levels of CDH1 and BMP1 reflect impairments in leukocyte adhesion and ECM homeostasis. Elevated levels of BATF3 and proC reflect altered immune cells recruitment and activation. Elevated levels of WNT5A reflect activation of non-canonical WNT signaling, leading to adipogenesis and inflammation. ADGRD1, G-protein coupled receptor D1; BATF3, basic leucine zipper transcriptional factor ATF-like 3; BMP1, bone morphogenetic protein 1; CDH1, cadherin-1; CDH5, Cadherin-5/VE-cadherin; CLEC3B, tetranectin; COMP, cartilage oligomeric matrix protein; CSF1, colony stimulating factor 1; DAG, diglyceride; ERVV1, endogenous retrovirus group V member 1; HTRA1, high-temperature requirement serine protease A1; KDR, kinase insert domain receptor; LRP8, low-density lipoprotein receptor-related protein 8; proC, Protein C; SELL, L-selectin; TAG, triglyceride; WNT5A, wingless-type MMTV integration site family, member 5A. **(C and D). Abnormalities in energy and lipid metabolism (C) before and (D) after exercise**. **C.** Before exercise, plasma levels of TAGs and DAGs were elevated in ME/CFS; ratios of ORN:CIT, and ARG: CIT, were elevated; levels of 12,13-diHOME were reduced. Levels of CIT were reduced in long duration ME/CFS. Reduced level of CIT, together with elevated ORN:CIT ratios, reflect impairments in urea cycle, leading to improper detoxification and inflammation. **D.** After exercise, plasma levels of citric acid increased in ME/CFS but decreased in HC; levels of CAR were reduced; ratios of ORN:CIT and ARG:CIT were decreased. Plasma levels of ASP increased in long duration ME/CFS. Increased levels of citrate reflect TCA cycle impairment, increased fatty acid synthesis, and inflammation. Decreased ratios of ORN:CIT and ARG:CIT, together with increased levels of ASP reflect impairments in urea cycle possibly due to reduced activity of CIT transporters or of enzymes for urea cycle intermediates. Increased levels of 12,13-diHOME and reduced levels of CAR reflect decreased flux of β-oxidation, further accumulating TAGs and DAGs. ARG, arginine; ASP, aspartic acid; CIT, citrulline; CAR, carnitine; DAG, diglyceride; diHOME, dihydroxyoctadecenoic acid; ORN, ornithine; TAG, triglyceride. **(E and F). Abnormalities in gut microbiome and TRP metabolism (E) before and (F) after exercise**. **E**. Before exercise, plasma levels of TFF1 and ILA were reduced in ME/CFS; levels of GA were elevated; ratios of KYN:TRP were reduced, ratios of KYNA:KYN were elevated. Plasma levels of CIT were reduced in long duration ME/CFS. Reduced levels of ILA and leucate reflect dysbiosis and enhanced inflammation in the gut. Elevated levels of GA reflect deconjugation of xenobiotic metabolites, leading to inflammation. **F.** After exercise, plasma levels of PPA and GA were elevated in ME/CFS; ratios of KYN:TRP were increased while ratios of KYNA:KYN were decreased. Plasma levels of REG4 were elevated in long duration ME/CFS. Increased ratios of KYN:TRP and increased plasma levels of 3-MA reflect activation of KYN pathway, diverting the TRP metabolism from serotonin pathway, leading to reduced serotonin production and cognitive dysfunction. Elevated levels of PPA reflect dysbiosis, leading to mitochondrial dysfunction and, potentially, neuroinflammation. Elevated levels of REG4 reflect compromised regeneration of gut lining, leading to impaired gut barrier function. CIT, citrulline; GA, glucuronic acid; HAAO, 3-hydroxyanthranilate 3,4-dioxygenase; IAA, Indole-3-acetic acid; ILA, indole-3-lactate/indole lactic acid; IPPA, indole-3-propionic acid; IPYA, indole-3-pyruvic acid; KYN, kynurenine; KYNA, kynurenic acid; PPA, propionic acid; QA, quinolinic acid; REG4, regenerating islet-derived 4; TRP, tryptophan; TFF1, Trefoil Factor 1; 3-HAA, 3-hydroxyanthranilic acid; 3-HK, 3-hydroxykynurenine; 3MA-3-methoxy anthranilate; 5-HT, 5-hydroxytryptamine.

Plasma multi-omics analyses indicated disturbances in mitochondria and dysregulation in TCA and FA pathways that may lead to fatigue and PEM (**Fig. 4C, 4D**). Increased levels of citrate following exercise suggest a lesion in the TCA cycle wherein acetyl-CoA combines with oxaloacetate to form citrate. Carnitine reduction after exercise impairs energy production via β-oxidation of FAs, further contributing to energy depletion and PEM. With increased levels of 12,13-diHOME and reduced levels of acylcarnitines, we postulate that there is inadequate oxidation of FAs in ME/CFS in response to exercise. Elevated post-exercise levels of GDF15, a biomarker for mitochondrial diseases, correlated with symptoms of fatigue and are consistent with PEM and may reflect systemic inflammation.^78^

Fatigue and PEM in ME/CFS may also include cognitive dysfunction, often referred to as “brain fog”. Studies in ME/CFS have reported altered plasma levels of amino acids critical to brain function, including glutamate, glutamine, tryptophan, serotonin, histidine, serine, taurine, and tyrosine.^46,58^ Branched-chain amino acids are also reported to be reduced in the urine and plasma in ME/CFS.^79^ We extended these findings and found impairments in tryptophan metabolism before and after exercise (**Fig. 4E, 4F**). Energy depletion in the CNS may lead to cognitive dysfunction, compounded by dysregulation in TRP-dependent pathways for the synthesis of KYN and serotonin.^80^ Post-exercise elevated levels of 3-methoxyanthranilate correlated with general fatigue in ME/CFS, highlighting the potential impact of KYN pathway on symptom severity (**Fig. 3B**). Dysregulation in neurotransmitter metabolism correlated with clinical symptoms (**Fig. 3A, 3B**).

Although the primary function of the urea cycle is ammonia detoxification, it also contributes to energy production by producing fumarate for the TCA cycle and NAD for ATP synthesis (**Supplementary Materials 2.3.9.2** for dysregulated NAD+ signaling). Dysregulated protein synthesis (EIF1) and accumulation of misfolded or aberrant proteins (UBE2D3) were associated with MFI scores of reduced activity and physical fatigue, respectively (**Fig. 3A**). As reflected in elevated after-exercise levels of CUTC and AOC2, disturbances in copper-dependent oxidant pathways associated with oxidative stress were correlated with general fatigue in ME/CFS (**Fig. 3B, 3D**). The persistent cellular stress, characterized by impairments in urea cycle, xenobiotic metabolism, and protein metabolism, may initiate response pathways that perpetuate mitochondrial dysfunction and immune dysregulation.

In addition to cytokine responses to SEB in TruCulture assays, subgroup-specific effects were observed in plasma metabolomic and proteomic analyses. For example, whereas female subjects with ME/CFS had amino acid abnormalities indicative of glutathione (GSH) depletion both before and after exercise (**Supplementary Materials 2.3.11** for abnormalities in methionine cycle and GSH biosynthesis), male ME/CFS had lower plasma levels of plasmalogens, suggesting peroxisomal dysfunction (**Supplementary Materials 2.3.10** for peroxisomal dysfunction).

Two recent publications support a role of innate immunity dysregulation in ME/CFS. A review from Humer et al. details evidence that the pathogenesis of ME/CFS entails trained immunity.^81^ In trained immunity, infection with one pathogen results in a reduced threshold for triggering inflammation by other pathogens. This model is congruent with results presented here, wherein ME/CFS patients have a lower threshold than healthy subjects for immune activation following exposure to the representative components of multiple microbes.

Consistent with our findings and previous evidence of redox imbalance in ME/CFS,^82^ Shankar et al. found evidence of cellular and systemic oxidative stress in PBMCs from both ME/CFS and long-COVID patients that result in inflammation and mitochondrial damage.^83^ They also reported sex-specific differences that were consistent with our findings in plasma, wherein female ME/CFS subjects had elevated reactive oxidative species (ROS) and depleted GSH, and male ME/CFS subjects had biomarkers for lipid peroxidation^83^. T cell proliferative responses in PBMC triggered by ROS were mitigated by metformin, a drug that reduces oxidative stress.^83^

In concert, work presented here and by these two independent research teams provides strong support for investing in research focused on dissecting innate immunity in ME/CFS. It also suggests potential candidates for clinical trials. In addition to metformin, the regulatory cytokine IL-37 and the mTOR inhibitor rapamycin may be helpful to ME/CFS individuals with evidence of enhanced innate immunity or hypersensitivity to microbial stimuli.^84^ Patients with evidence of dysbiosis may benefit from dietary supplementation with prebiotics (inulin) and probiotics (*F. prausnitzii*) that enhance GI barrier integrity and regulate immune responses.^50^ Low baseline levels of 12,13-diHOME and high post-exercise levels of GDF15 may identify individuals with pronounced metabolic disruption, more likely to respond to dietary supplementation with 12,13-diHOME or to treatment with GDF15-neutralizing antibody.^85^ Individuals with abnormalities in tryptophan metabolism, may respond to supplementation with 5-hydroxytryptophan or selective serotonin reuptake inhibitors.^86^ In patients with low carnitine levels, carnitine supplementation may restore carnitine shuttle function and enable use of lipids for energy source. Estrogen supplements may modulate the inflammatory response in older women with ME/CFS. Although the pathogenesis of ME/CFS remains obscure, mechanistic dissection of its pathobiology is revealing subtype-specific biomarkers that may inform clinical research and lead to targeted interventions.

### Limitations

This study is limited by the use of plasma samples for metabolomic and proteomic analyses. Testing mechanisms that provide explanations for the direction and magnitude of metabolic disruptions require isotope-labelled metabolic flux studies in specific cells, organoids or tissues. Cells from muscle and other metabolically active organs are not available from ME/CFS patients; and animal models with bona fide ME/CFS symptoms have not been established. Our findings provide an overview of systemic changes but may not accurately capture cell- or tissue-specific abnormalities. In addition, small sample sizes in the subgroup analyses, as well as in the dose- and time-dependent cytokine analyses in PBMCs in response to LPS and poly I:C, may affect the generalizability of the findings. Nonetheless, results from metabolomic, proteomic, and TruCulture analyses were convergent and consistent with ME/CFS symptoms. Lastly, although a range of important covariates were adjusted for in our study, we cannot rule out diet, medications, and lifestyle as potential confounding factors.

## Methods

### Clinical assessments, cardiopulmonary Exercise (CPX) testing, and plasma collection

Clinical symptoms and health status of both ME/CFS and HC were assessed at 3 time points: pre-exercise (same day), 24 hours, and 48 hours post-exercise. The assessments used the following instruments: the MFI, the Short Form 36 Health Survey (SF-36), DePaul Symptom Questionnaire (DSQ), and Pittsburgh Sleep Quality Index (PSQI) (**Supplementary Materials 1.1** for clinical assessments).

Each participant performed a cardiopulmonary exercise test on an upright ergocycle, using an individualized ramp-based protocol as per consensus guidelines.^87^ Continuous electrocardiogram monitoring was performed, and CPX variables were documented breath-by-breath. The blood pressure was measured at regular intervals during the study on the arm opposite to the one with the intravenous cannula. Participants were encouraged to exercise to their maximal capacity; however, the test was terminated upon the participant’s request or if abnormal electrocardiographic findings were observed.

Blood samples were collected at two time points: pre-exercise (same day) and 24 hours post-exercise. A 12-hour fasting period was required before each blood draw. Whole blood collected in BD VacutainerTM Cell Preparation Tubes (CPT) were shipped to Columbia University at 4°C and processed as per manufacturer’s protocol. The plasma layer was transferred into tubes without disturbing the membrane and centrifuged at 810 Relative Centrifugal Force (RCF) for 10 minutes. The supernatants were stored at -80°C as aliquots of plasma.

### *Ex-vivo* blood stimulation and cytokine measurements

Blood samples were collected into TruCulture tubes (Myriad) prepared with null (no stimulant), LPS, SEB, poly I:C, or HKCA.^7^ The tubes were then placed in a dry block incubator and maintained at 37°C for 48 hours. After incubation, valves were inserted to separate cells from the supernatant. Supernatants were aliquoted and were kept frozen at −80°C prior to further analyses.

TruCulture OptiMAPTM assay (Myriad RBM) was used to analyze the concentrations of cytokines ENA-78 (CXCL5), GM-CSF, IFN-γ, IL-1β, IL-10, IL-12p70, IL-13, IL-17, IL-2, IL-23, IL-6, IL-8, and TNF-α. All samples were run in duplicate alongside the calibration standards and controls in the same assay plates. The assay employs antigen-specific antibodies that are optimized in a capture-sandwich format. The mean fluorescence intensities of analyte-specific beads were estimated using the Luminex platform.^7^ To control for lot-to-lot variation, calibration standards were adjusted via standard curve-fitting to match prior lot concentrations.

### *In-vitro* PBMC stimulation with LPS and poly I:C

The PBMCs from pre-exercise ME/CFS (n=8) and controls (n=7) were thawed and seeded at density of 100,000 cells per well in 12 well cell culture plates. We used LPS (E. coli O55: B5, Sigma) and poly I:C (Sigma, high molecular weight) for stimulation of stored PBMCs. We used 0.01, 0.1 and 100 ng/ml of LPS and 0.2, 2 and 20 µg/ml of poly I:C for 48 hours and incubated at 37°C with 5% CO_2_. Samples were collected at 12, 24 and 48 hours of incubation and measured for induction of IL-6 using Luminex xMAP based system ProcartaPlex (Invitrogen) following manufacturer’s protocol. The data were normalized using standard curve and plotted for IL-6.

### Plasma metabolomics and external validation

Untargeted metabolomics data were acquired using three chromatography/ mass spectrometry-based (MS) assays into primary metabolites (PM), biogenic amines (BA), and complex lipids, while targeted metabolomics was used to acquire oxylipins (OL) data. The details for each panel and subsequent standardized data processing are described in **Supplementary Materials 1.3 & 1.4**.

Data for external validation of metabolomic analysis were obtained from a cohort reported by Germain et al. (2022) that included plasma levels of 933 known metabolites in 60 ME/CFS patients (45 female, 15 male) and 45 matched HC subjects (30 female, 15 male) before and after 2 maximal exercise test challenges separated by 24 hours^88^. Our pre- and 24 hours post-exercise time points corresponded to before exercise on day 1 (D1PRE) and before exercise on day 2 (D2PRE), respectively, in the validation cohort. A total of 220 metabolites were matched between the two studies, including 85 PMs, 111 BAs, 18 CLs, and 5 OLs. Validation for plasma metabolomic analysis was conducted on the biological pathway level using MetaboAnalyst 6.0,^89^ identifying significant pathways (*p*<0.1) comparing ME/CFS vs. HC before and 24 hours after exercise (results are reported in **Supplementary Materials 2.5, Table S10**). A total of 566 metabolites from our study and 562 metabolites from the external validation study were annotated in MetaboAnalyst.

### Plasma proteomic assays

Plasma proteomic analysis was performed using the SomaScan Assay v4.1 developed by SomaLogic, Inc.. The platform quantifies sensitive and highly reproducible proteomic data for over 7000 proteins using DNA aptamers that bind to target proteins. In each plate, pooled reference standards and buffer standards were included to control for possible batch effect. Samples normalization was performed using median signal intensities within and between plates for reference standards, which controlled for technical variations. The proteomic assay included 7,285 proteins with no missing values. After excluding protein analytes that did not pass the quality control criteria, we analyzed data from 6,969 proteins.

### Statistical Analyses

Statistical analyses were performed using R version 4.3.2 (RStudio, Inc., Boston, MA) and MATLAB and Statistics Toolbox R2021a (MathWorks, Inc., Natick, MA). All *p*-values were 2-tailed. The details of data pre-processing and outlier exclusion are included in **Supplementary Materials 1.4**.

### Regression analysis

Given the multi-level clustered structure in our data (matched case-control pairs and within-subject repeated measures), we applied linear mixed-effect models (LMMs) to compare the levels of each molecular analyte between groups and within subjects. The models were adjusted for age, sex, race/ethnicity, geographic/clinical site, BMI, and self-reported irritable bowel syndrome (sr-IBS). The fixed effects included ME/CFS status (case vs. HC), time point (post-exercise vs. pre-exercise), and their interaction term. Besides the marginal intercept, we included two random intercepts: one accounting for the matched pair correlations and one accounting for within-subject correlations. The unmatched subjects were also included in the LMMs only accounting for the within-subject correlations. Inferences on five comparisons were derived from LMMs: 1) differences in analyte levels between ME/CFS and HC before exercise, 2) differences in analyte levels between ME/CFS and HC after exercise, 3) changes in analyte levels in ME/CFS from pre- to post-exercise, 4) changes in analyte levels in HC from pre- to post-exercise, and 5) differences in analyte trajectories between ME/CFS and HC, reflecting interaction/crossover effects. To facilitate interpretation, we used “higher/lower”, “elevated/reduced”, or “accumulated/depleted” to describe significant between-group differences, i.e., in comparisons 1) and 2); We used “increased/decreased” to specifically describe significant within-group changes or significant crossovers, i.e., in comparisons 3), 4), and 5). Multiple comparisons in each molecular assay were corrected using the Benjamini-Hochberg procedure^90^ controlling the false discovery rate (FDR) at the 0.05 level.

### Bayesian Analysis

Bayesian alternatives to the null hypothesis significance testing (NHST) framework have been shown to improve the biological interpretability of metabolomic data from human cohorts.^91^ For each molecular analyte, we conducted the LMMs paired with Bayesian analysis with weakly informative priors (**Supplementary Materials 1.5** for details in Bayesian analyses), yielding BayesFactors (BFs) and 95% highest density credible intervals (HDIs). We considered a comparison to be significant if BF>3 and 95% HDI does not cover 0, or if the FDR adjusted *p*-value<0.05.

Bayesian statistics are most powerful when previous knowledge is incorporated into the models as “prior information”. This strategy was adopted in our metabolomics analysis using data from an external validation cohort. For each of the 220 matched metabolites, we used the results from Germain et al. (2022)^88^ as prior distribution and derived combined BFs (BF_combined_) and HDIs (HDI_combined_). For these metabolic analytes, a significant comparison was identified, besides the criteria listed above, if BF_combined_>10 and HDI_combined_ does not cover 0.

### Enrichment analyses

For the metabolomic analysis, we performed chemical enrichment analysis using ChemRICH^92^ to determine chemical classes that were significantly altered. We also applied the Ingenuity Pathway Analysis (IPA version 11172566; QIAGEN) integrating the metabolomic and the proteomic datasets. The significant criteria listed above were incorporated into both ChemRICH and IPA analyses.

### Analyte-symptom correlation analyses

We investigated whether the levels of metabolomic and proteomic analytes, that showed significant associations with ME/CFS pre- or post-exercise, correlated with the MFI symptom severity scales at the corresponding time point in ME/CFS. Drug metabolites were excluded from this analysis. We employed regularized canonical correlation analysis (RCCA) with Lasso penalty to avoid potential overfitting. Full data are shown in **Table S9**. Details of analyte-analyte correlations are described in **Supplementary Materials 1.6**.

## Data Availability

All data and analysis code are uploaded to RTI International, Data Management Coordinating Center (DMCC) for MECFS Research Network (ME/CFSnet). Metabolomics data are also uploaded to the data repository at Metabolomics Workbench.

## Acknowledgements

The authors are grateful for generous support from Hutchins Family Foundation through the Chronic Fatigue Initiative, NIH/NIAID grant # 4U54AI138370-06 and of AJ from the Ansell Family Foundation.

