## Supplement material for "Heightened innate immunity may trigger chronic inflammation, fatigue and post-exertional malaise in ME/CFS"

**Supplementary Materials**

1. **Supplementary Methods**
   1.1. **Clinical assessments.**

Multidimensional Fatigue Inventory (MFI) comprises of a 20-item self-reported questionnaire focused on general, physical and mental fatigue, reduced activity, and reduced motivation.^1^ The SF-36 includes the following subject-reported evaluations about current health status: physical and social functioning, physical and emotional limitations, vitality, pain, general health perceptions, and mental health status.^2^ Self-reported cognitive function was obtained from the DSQ^3^ questionnaire data and was scored using a standard cognitive disturbance definition as well as a modified definition based on a subset of questionnaire variables. Sleeping disturbances linked to ME/CFS were tested and scored based on DSQ and PSQI questionnaire items^3,4^. Each instrument was transformed into a 0–100 scale to facilitate combination and comparison wherein a score of 100 is equivalent to maximum disability or severity and a score of zero is equivalent to no disability or disturbance. Correspondingly, the SF-36 scores were reversely coded after the original scales were proportionately expanded to a 0-100 scale. A self-reported diagnosis of irritable bowel syndrome (sr-IBS) was based on answers in the medical history form. Subjects were asked if they had received a previous IBS diagnosis by a physician and the date of that diagnosis.

1.2. **Subgroup stratifications**.

Factors commonly associated with group-specific findings in ME/CFS include sex,^5,6^ age,^7^ and duration of illness.^8,9^ We conducted corresponding stratified analyses. For age-stratification, we divided the female cohort into subjects younger or older than 45 years of age (YoA).^10^ This cutoff was chosen based on the observation that in the U.S., the average age for menopause is 51 YoA, with perimenopause typically occurring 4-5 years prior. Age-stratification was not performed in males due to limited sample size (**Table 1**). For illness duration, the cutoff was set at 3 years.^8,11^ Subgroup comparisons are presented as: female subjects with ME/CFS (**ME/CFS^F^**) vs. female HC subjects (**HC^F^**); male subjects with ME/CFS (**ME/CFS^M^**) vs. male HC subjects (**HC^M^**); female subjects with ME/CFS <45 years of age (YoA) (**ME/CFS^YF^**) vs. female HC subjects <45 YoA (**HC^YF^**); female subjects with ME/CFS >45 YoA (**ME/CFS^OF^**) vs. female HC subjects >45 YoA (**HC^OF^**); ME/CFS subjects with short duration (<3 years) of illness (**ME/CFS^SD^**) vs. **HC**; ME/CFS subjects with long duration (>3 years) of illness (**ME/CFS^LD^**) vs. **HC**.

Plasma metabolomic and TruCulture cytokine analyses included all study subjects. Plasma proteomic analyses included all subjects from the NY site (**Table 1**). ME/CFS cases and HC were similar for the matching variables (sex, age, race/ethnicity, geographic site). In the plasma metabolomic and TruCulture cytokine analyses, sex stratification yielded 37 ME/CFS^F^ subjects, 37 HC^F^ subjects, 19 ME/CFS^M^ subjects, and 15 HC^M^ subjects; age stratification in females yielded 18 ME/CFS^YF^ subjects, 12 HC^YF^ subjects, 19 ME/CFS^OF^ subjects, and 25 HC^OF^ subjects; illness duration stratification yielded 13 ME/CFS^SD^ subjects and 42 ME/CFS^LD^ subjects. In the plasma proteomic analysis, sex stratification yielded 24 ME/CFS^F^ subjects, 25 HC^F^ subjects, 12 ME/CFS^M^ subjects, and 9 HC^M^ subjects; age stratification in females yielded 9 ME/CFS^YF^ subjects, 7 HC^YF^ subjects, 15 ME/CFS^OF^ subjects, and 18 HC^OF^ subjects; illness duration stratification yielded 12 ME/CFS^SD^ subjects and 23 ME/CFS^LD^ subjects.

1.3. **Metabolomic assays.**

Primary metabolites (PM) such as mono- and disaccharides, hydroxyl- and amino acids were measured by gas chromatography/time-of-flight mass spectrometry (GC-TOF MS)^12^ including data alignment and compound annotation using the BinBase database algorithm.^13^ Biogenic amines (BA) including microbial compounds such as TMAO, methylated and acetylated amino acids and short di- and tripeptides were measured by hydrophilic interaction liquid chromatography/quadrupole time-of-flight mass spectrometry (HILIC-QTOF MS). Complex lipids (CL) including phosphoglycerolipids, triacylglycerides, sphingolipids and free fatty acids were analyzed by liquid chromatography (LC)/quadrupole time-of-flight mass spectrometry (CSH-QTOF MS).^14^ Targeted bioactive oxylipin (OL) assay included thromboxanes, prostaglandins, and hdyroxy-, keto- and epoxy-lipins. All LC-MS/MS data included diverse sets of internal standards. LC-MS data were processed by MS-DIAL vs. 4.0 software,^15^ and the compounds were annotated based on accurate mass, retention time and MS/MS fragment matching using LipidBlast^16^ and Massbank of North America libraries.^17^ MS-FLO was used to remove erroneous peaks and reduce the false discovery rate in LC datasets.^18^ Some compounds were detected in multiple assays, and certain complex lipids were annotated as different structural isomers or were detected in both positive (ESI+) and negative (ESI-) ion modes, resulting in a total of 935 metabolic analyte. Data were normalized by SERRF.^19^ Residual technical errors were assessed by coefficients of variation (CV) for known metabolites.

1.4. **Data pre-processing.**

Twenty two of the 935 metabolomic analytes were excluded from the statistical analysis because more than 50% of the samples had missing values, reflecting measurements below the detection/quantification limits (**Table S2A**). Most of the excluded metabolites were pharmacotherapeutic compounds. For each of the remaining metabolic analyte, missing values were replaced with half of its smallest available value.^20,21^ There were no missing values in the proteomics dataset. In the TruCulture cytokine analysis, GM-CSF (100%), IFN-γ (99.1%), IL-1β (98.1%), IL-2 (100%), IL-6 (99.5%), IL-10 (100%), IL-12p70 (100%), IL-17 (99.5%), IL-23 (100%), and TNF-α (96.8%) were excluded in the no stimulant panel; GM-CSF (87.1%), IL-2 (87.6%), IL-10 (76.1%), IL-13 (59.2%), IL-17 (71.5%), and IL-23 (90.8%) were excluded in poly I:C; IL-12p70 (99.1%) and IL-23 (53.2%) were excluded in HKCA; IL-12p70 (50.9%) was excluded in LPS; no analytes were excluded in SEB. Missing values below the lower limit of quantitation (LLOQ) in each of the remaining cytokines were replaced by half of its LLOQ value. Certain cytokines in the TruCulture analysis had levels above the upper limit of quantitation (ULOQ), and they included: in HKCA, CXCL5 (n=1), and IL-8 (46.8%); in SEB, IL-8 (n=5). Due to the small numbers, the semi-quantitative values of CXCL5 in HKCA and of IL-8 in SEB were replaced with the corresponding ULOQ values. Levels of IL-8 in HKCA were analyzed both continuously (replacing >ULOQ with the ULOQ value) and dichotomously (>ULOQ vs. <ULOQ).

Outliers were identified using principal component analysis (PCA) in each assay, separately (**Table S2B**). After the outliers were removed, levels of each analyte were natural log transformed.

1.5. **Bayesian analyses.**

For each molecular analyte, we conducted Bayesian analysis on the LMM regression models using R packages “rstanarm” ^22^and “bayestestR”.^23^ Default (weakly informative) prior distributions were applied adjusting the scales of the priors internally. We then calculated the BayesFactors (BFs) and 95% highest density credible intervals (HDIs). BFs are ratios that quantify the probability of the alternative hypothesis (β≠0) over the null hypothesis (β=0) by estimating the strength of evidence.^24^ The 95% credible intervals are a range of values within which the true effect falls at 95% confidence, given the data.^25,26^

1.6. **Analyte-analyte correlation analyses.**

We investigated differences in the inter-analyte relationships between ME/CFS and HC, by building an analyte-analyte correlation network at each time point using all analytes significantly associated with ME/CFS. Drug metabolites were excluded. Spearman correlation paired with Bayesian statistics was performed between each pair of significant analytes, and the correlations with BF>3 were included in the networks. Differences between the inter-analyte networks in ME/CFS and HC were inferred using R package “igraph”. We assessed modules within network identified using the Louvain Method.^27^ Within each module, the center, or the module hub, was determined using eigenvector centrality score.

2. **Supplementary Results and Discussion**
   2.1. **Study population characteristics.**

All ME/CFS subjects completed standardized screening and assessment instruments including medical history and symptom rating scales as well as a physical examination. Healthy controls (HC) underwent the same screening process as ME/CFS subjects and were not recruited if they reported ME/CFS or other conditions deemed by the recruiting physician to be inconsistent with HC status. In addition, subjects (ME/CFS and HC) were not recruited if they were immunosuppressed, had a history of alcohol or substance abuse, psychiatric illness, antibiotics in the prior six weeks, or clinically significant findings on physical exam or screening laboratory tests.

The comorbidity of self-reported irritable bowel syndrome (sr-IBS) was more prevalent in ME/CFS cases than in HC (Chi-squared, *p*=0.016). ME/CFS cases and HC differed in the MFI scales.^1^ We proportionately expanded the MFI original scores to 0-100, where 0 = no disability or disturbance and 100 = maximum disability or severity. The between-group differences in the MFI scores were significant at each time point: before, and 24 and 48 hours after exercise (Wilcoxon rank-sum, *p*<0.001). ME/CFS patients had more severe MFI general fatigue scores 48 hours after exercise than before and 24 hours after exercise (Wilcoxon signed-rank, *p*<0.001).

All study subjects performed a cardiopulmonary exercise test. Similar to the reports from other investigators,^28,29^ peak VO2 was significantly lower in ME/CFS than in HC: 1474 ± 463 vs. 1849 ± 639 ml/min, or 20.6 ± 5.9 vs. 27.5 ± 8.3 ml/kg/min (both *p* < 0.01).

Results from subgroup comparisons are presented as: **ME/CFS^F^** vs. **HC^F^** (**Fig. S2**); **ME/CFS^M^** vs. **HC^M^** (**Fig. S3**); **ME/CFS^YF^** vs. **HC^YF^** (**Fig. S4**); **ME/CFS^OF^** vs. **HC^OF^** (**Fig. S5**); **ME/CFS^SD^** vs. **HC** (**Fig. S6**); **ME/CFS^LD^** vs. **HC** (**Fig. S7**).

2.2. **Analytical datasets.**

In the metabolomic assays, targeted and untargeted mass spectrometry yielded data for 935 metabolites. We excluded 22 metabolites from statistical analyses because more than 50% of samples had measurements below the limits of detection (**Table S2A**). The remaining 913 metabolites comprised 143 primary metabolites (PM), 256 biogenic amines (BA), 445 complex lipids (CL), and 69 bioactive oxylipins (OL). We also acquired metabolomic data from an external validation cohort,^30^ wherein a total of 220 metabolites were matched between the two studies, including 85 PMs, 111 BAs, 18 CLs, and 5 OLs. Plasma proteomic analyses included 7,285 proteins with no missing values. After excluding analytes that did not pass the quality control criteria, we analyzed data from 6,969 proteins. In the TruCulture immunological assays, cytokines with more than 50% values below the lower limit of quantitation (LLOQ) were excluded, and they included GM-CS, IFN-γ, IL-1β, IL-2, IL-6, IL-10, IL-12p70, IL-17, IL-23, and TNF-α in the no stimulant panel; GM-CSF, IL-2, IL-10, IL-13, IL-17, and IL-23 in polyinosinic:polycytidylic acid (poly I:C); IL-12p70 and IL-23 in heat-killed *Candida albicans* (HKCA); IL-12p70 in lipopolysaccharide (LPS). The data characteristics, including median, interquartile range, and min-max range, of analytes in all assays at pre- and post-exercise time points are summarized in **Table S1**. Metabolomic data from the external validation cohort^30^ was also used as prior information in the Bayesian analyses.^31^

2.3. **Plasma metabolomic and proteomic analyses in subgroups of ME/CFS.**
     2.3.1. **Proteomic correlates of impaired pathogen recognition in ME/CFS^YF^.**

Exposure to circulating bacterial LPS, persistent viral infections, and viral reactivations are often reported in ME/CFS.^32-35^ Here, we report evidence for impaired pathogen recognition in ME/CFS^YF^. Integrative IPA analysis identified altered pathways of pattern recognition receptors (PRRs) in recognition of bacteria and viruses and macrophage activation in ME/CFS^YF^ both before and after exercise (**Fig. S4C, Table S7D**). Analytes associated with TLR4 cascades exhibited a significant interaction effect (**Table S7D**).

Before exercise, plasma levels of TLR3, Dectin-1 (CLEC7A), and human leukocyte antigen (HLA)-DR β3 (HLA-DRB3) were reduced in ME/CFS^YF^ (**Fig. S4B**). Levels of Dectin-1 were also lower after exercise (**Fig. S4B**). TLR3 binds to double-stranded RNA ligands within endosomes, eliciting immune responses against viral pathogens.^36^ Dectin-1 recognizes β-glucans and initiates immune responses against fungal and certain bacterial pathogens.^37^ Besides activating complement and coagulation systems, HLA-DRs are major histocompatibility complex (MHC) class II (MHCII) proteins critically involved in antigen presentation and subsequent T cell activation.^38^ Recent studies suggest that lower expression of HLA-DR proteins indicates reduced functional capacity of antigen-presenting cells (APCs) in inflammatory diseases such as sepsis and acute pancreatitis.^39,40^ Lower levels of TLR3, Dectin-1, and HLA-DRB3 are consistent with inadequate basal immune activation capabilities in ME/CFS^YF^, that limits the formation of early endosomes and phagosomes necessary for the recognition and clearance of viral and other intracellular pathogens.

Following exercise, TLR4|LY96 had an increased interaction effect, wherein levels were increased in ME/CFS^YF^ but decreased in HC^YF^ (**Table S6D**; the assay did not distinguish TLR4 from its co-receptor). Levels of TRAM (TIR domain-containing adapter molecule 2, TICAM2) were elevated in ME/CFS^YF^ after exercise (**Fig. S4B**). TLR4 is a key component of innate immunity against bacterial LPS and mediates the production of pro-inflammatory cytokines via MyD88-dependent or TRIF-dependent pathways.^41-43^ In the TRIF-dependent pathway, adapter protein TRAM recruits TRIF to activate TLR4 signaling independent of MyD88.^44^ MyD88 and TRIF were not included in the proteomic assay; however, we measured serine/threonine-protein kinase TBK1 which is critically involved in both pathways.^45,46^ In contrast to TLR4, levels of TBK1 were decreased in ME/CFS^YF^ but increased in HC^YF^ in response to exercise, and the post-exercise levels were lower in ME/CFS^YF^ (**Table S6D**). TBK1 is essential for inducing type I interferon signaling through interferon regulatory factor 3 (IRF3).^47^ Post-exercise levels of IRF3 in ME/CFS^YF^ exhibited a trend towards reduction (**Table S6D**, *p*=0.030, BF=1.062). Despite increased levels of TLR4 and higher levels of TRAM, reduced levels of TBK1 and IRF3 indicate inadequate activation of both MyD88- and TRIF-dependent pathways, which leads to compromised pathogen clearance. Moreover, the MyD88-TBK1 pathway promotes glycolysis.^45^ We observed evidence of inhibited glycolysis in ME/CFS^YF^ following exercise (see **Supplementary section 2.3.8.1** for mitochondrial dysfunction and lipid abnormalities in ME/CFS^YF^). TBK1 also acts as an anti-inflammatory factor in macrophages, suppressing NF-κB and MAPK pathways and IL-1β production.^48^ Decreased levels of TBK1 may contribute to inflammation. These observations provide evidence indicating impairements in PRRs both before and after exercise in ME/CFS^YF^ that may lead to inefficient clearance of pathogens and consequent inflammation.

2.3.2. **Elevated type I interferon signaling after exercise in ME/CFS^YF^ and ME/CFS^OF^.**

Despite impaired pathogen recognition (TLR3/4 signaling) in ME/CFS^YF^, elevation of type I interferon signaling was observed after exercise in both ME/CFS^YF^ and ME/CFS^OF^, indicating enhanced interferon-mediated inflammation and PEM. After exercise, interferon regulatory factor (IRF) activation by pattern recognition receptors (PRRs) pathway in ME/CFS^YF^ and interferon signaling pathway in ME/CFS^OF^ were significantly altered (**Fig. S5C, Table S7D**).

In ME/CFS^YF^, post-exercise levels of type I interferon IFNA10 were higher (**Fig. S4B**). IFN-α is mainly produced by plasmacytoid dendritic cells and mediates antiviral responses.^49^ Type I interferons bind to receptors IFNAR1 and IFNAR2, thereby activating canonical pathways via JAK/STAT and non-canonical pathways via MAPK and PI3K/AKT/mTOR.^49,50^ IFN-α upregulates the expression of immune checkpoint protein PD-L1 (CD274) on dendritic cells through STAT3 and p38 MAPK pathways.^51^ In Th17 cells, IL-21 is induced with high promoter binding activity by STAT3.^52^ Consistent with higher IFN-α levels, post-exercise levels of PD-L1, IL-21, and IL-6 were elevated in ME/CFS^YF^ (**Fig. S4B**). However, we did not observe differences in PD-1 levels (**Table S6D**).

In ME/CFS^OF^, post-exercise levels of transcription factor IRF1 were elevated (**Fig. S5B**). IRF1 is induced in response to infection and regulates gene expression central for immune response.^53^ It mediates the activation of PRRs, including TLR4, to enable pathogen clearannce.^53,54^ In macrophages, IRF1 promotes induction and accessibility of interferon-stimulated genes (ISGs), such as *MX1* and *OAS1*.^55^ IRF1 also facilitates the interferon-induced production of IL-15.^56^ IL-15 is involved in the protective immune response to microbial components through regulation of both innate and adaptive immunity.^57^ In line with higher levels of IRF1, post-exercise levels of MX1 and IL-15 were elevated in ME/CFS^OF^ (**Fig. S5B**). Levels of MX1 correlated with MFI scores indicating reduced motivation (**Fig. S5D**).

2.3.3. **Differences in NK cell dysregulation between ME/CFS^YF^ and ME/CFS^OF^.**

NK cells are important mediators of innate immunity, responsible for the clearance of infected and transformed cells.^58^ Reduced NK cell signaling has been documented in ME/CFS.^59^ We found that levels of analytes in NK cell signaling pathway were mostly elevated in ME/CFS^YF^ before exercise and were reduced in ME/CFS^OF^ both before and after exercise (**Fig. S5C, Table S7D & S7E**). Additionally, ME/CFS^YF^ had an increased interaction effect in NK cell signaling (**Table S7D**).

In ME/CFS^YF^, pre- and post-exercise levels of killer cell lectin-like receptor KLRB1 (CD161) were elevated (**Fig. S4B**). KLRB1 is a surface receptor on NK cells that facilitates NK cell maturation but also inhibits cytotoxicity through interactions with lectin-like transcript 1.^60^ In response to exercise, plasma levels of CD48, MHCI chain-related proteins MICA and MICB, and natural cytotoxicity triggering receptor NCR1 were increased in ME/CFS^YF^ but decreased in HC^YF^ (**Table S6D**). CD48 is the receptor for signaling lymphocyte activation molecule (SLAM) 2B4 (CD244), triggering NK cell activation.^61^ MICA and MICB are stress-induced ligands for NK cell activating receptor NKG2D, triggering effector functions in both NK and T cells.^62-64^ NCR1, another surface molecule on NK cells, initiates NK cell activation and effector functions.^65^ Studies have reported that, in healthy subjects, NK cell population and cytotoxicity increase during and immediately after maximal exercise, but drop below baseline 1-6 hours post-exercise.^66,67^ Increased levels of CD48, MICA, MICB, and NCR1 in ME/CFS^YF^ indicate prolonged NK cell activation and cytotoxicity after exercise, that may exacerbate inflammation in response to IFN-α (see **Supplementary Materials 2.3.2** for elevated type I interferon signaling in ME/CFS^F^).^68^

In ME/CFS^OF^, baseline levels of CD244 (high affinity ligand for CD48), MICA, KLRB1, and IgG Fc receptor FCGR3A were reduced (**Fig. S5B**). Levels of MICA and FCGR3A were also lower after exercise (**Fig. S5B**). KLRB1 reduction may lead to accumulation of immature NK cells.^69^ FCGR3A mediates the IgG effector function of NK cells, initiating antibody-dependent cellular cytotoxicity (ADCC).^70^ These findings in ME/CFS^OF^ may reflect the phenomenon of immunosenescence as NK cells are the primary drivers of immunosurveillance targeting senescent cells.^71^ Furthermore, ME/CFS^OF^ had lower levels of lactotransferrin (LTF) after exercise and lower levels of cytoplasmic dynein light chain 1 (DYNLL1) before and after exercise (**Fig. S5B**). Reduced post-exercise levels of LTF correlated with MFI reduced activity scores (**Fig. S5D**). Both LTF and DYNLL1 have been shown to play roles in NK cell cytotoxicity.^72-74^

2.3.4. **Additional innate immunity abnormalities in ME/CFS^F^.**

In ME/CFS^F^, the phagosome formation pathway was altered before and after exercise (**Fig. S2C**). Particularly, levels of FCGR1A (CD64) were reduced (**Fig. S2B**). FCGR1A is primarily produced by non-lymphoid cells and mediates the clearance of immune complexes.^75-77^

In ME/CFS^YF^, pathways associated with neutrophil degranulation and neutrophil extracellular trap (NET) signaling were altered before and after exercise; pathway for phagosome formation was altered before exercise (**Fig. S4C, Table S7D**). Post-exercise levels of CD47 were lower in ME/CFS^YF^ (**Fig. S4B**). CD47 maintains immune tolerance by inhibiting macrophage-mediated phagocytosis.^78^ It also facilitates cell-cell adhesion.^79^ Levels of sialic acid-binding Ig-like lectins SIGLEC5 and SIGLEC14 were elevated in ME/CFS^YF^ before and after exercise (**Fig. S4B**). SIGLEC5 and SIGLEC14 share high sequence similarity, tissue distributions, and ligand specificities, however, they have opposite functions and are, therefore, described as “paired receptors”.^80^ SIGLEC5 is upregulated upon LPS stimulation and, through interactions with β-protein, inhibits phagocytosis in macrophages and neutrophils, which can be rescued by SIGLEC14.^81-83^ Elevated levels of these “paired receptors” and reduced levels of CD47 in ME/CFS^YF^ reflect dysregulated phagocytosis, characterized by concurrent immune activation and modulation.

In ME/CFS^OF^, we observed abnormalities in MHCI-associated activities, as reflected by higher levels of inhibitory cell surface receptor LILRB2 both before and after exercise (**Fig. S5B**). LILRB2 is mainly expressed on myelomonocytic cells and binds to MHCIs in regulation of immune response.^84,85^ In monocytes, LILRB2 inhibits FCGR1A- and FCGR3A-mediated antibody-dependent cellular cytotoxicity (ADCC), accompanied by impaired calcium mobilization.^86^ LILRB2 elevation is consistent with FCGR1A depletion in ME/CFS^F^ and FCGR3A depletion in ME/CFS^OF^. LILRB2 is also a receptor for Aβ oligomers in the human brain and is posited to contribute to synaptic loss and cognitive impairment in AD patients.^87^

2.3.5. **Disrupted ECM homeostasis and dysregulated cell-cell adhesion in ME/CFS subgroups.**

In addition to the differential pathways and analytes discussed in cell-cell and cell-ECM interactions in main manuscript, in ME/CFS^F^, baseline levels of P-selectin (SELP) were reduced, and levels of lumican (LUM) and secreted protein acidic and rich in cysteine (SPARC, osteonectin) were reduced before and after exercise (**Fig. S2B**). SELP is expressed on the surface of endothelial cells and activated platelets and mediates recruitment of leukocytes during inflammation.^88,89^ LUM is a proteoglycan that plays critical roles in collagen assembly and ECM regulation.^90,91^ LUM binds with LPS and promotes the innate immune response through CD14-TLR4 pathway.^92^ LUM also facilitates neutrophil migration to sites of inflammation through binding to β2 integrin.^93^ Osteonectin is a calcium- and collagen-binding matricellular myokine that is secreted into blood after exercise and regulates cell-ECM interactions.^94^

In ME/CFS^M^, pathways involving integrin cell surface interactions and signaling of PIP3/AKT and FGFR were altered before exercise (**Fig. S3C, Table S7C**). Baseline levels of Grb2-associated binder 1 (GAB1) were lower (**Fig. S3B**). GAB1 is an insulin receptor substrate 1-like adapter protein involved in assembling functional complexes downstream of receptor tyrosine kinases (RTKs) and plays a critical role in regulating inflammation.^95-97^ It is also recruited by β1 integrin to mediate cell adhesion.^98^

In ME/CFS^YF^, baseline levels of intercellular adhesion molecules ICAM1, ICAM2, and ICAM4 were lower (**Fig. S4B**). ICAM4 levels were also lower after exercise (**Fig. S4B**). Although not significant, post-exercise levels of ICAM1 and ICAM2 trended toward reduction (**Table S6D**). Following selectin-mediated leukocyte rolling, ICAM1 and ICAM2 interact with β2-integrin and facilitate the firm adhesion, recruitment, and migration of leukocytes to sites of inflammation.^99,100^ ICAM4 is erythroid-specific and promotes erythrocyte-platelet adhesion.^101^ ICAM1 also stimulates T cell activation, induces NK cell-mediated cytotoxicity, and maintains barrier integrity at endothelial and epithelial regions.^102-104^ Plasma levels of CD63 were elevated in ME/CFS^YF^ both before and after exercise (**Fig. S4B**). Tetraspanin family member CD63 is an endosomal resident protein with pleiotropic functions, including platelet activation, neutrophil activation and degranulation, and cell-ECM adhesion.^105-107^ CD63 maintains ECM homeostasis; it is functionally involved in E2/C5 cell adhesion to the ECM.^107,108^ In addition, CD63 interacts with metalloproteinase inhibitor 1 (TIMP1), which inhibits matrix metalloproteinase (MMP) enzymatic activity.^109^ Taken together, reduced levels of ICAMs and elevated levels of CD63 demonstrate additional insights into the mechanisms of dysregulated cellular adhesion and disrupted ECM homeostasis, consistent with those in ME/CFS, ME/CFS^F^, and ME/CFS^M^.

ME/CFS^SD^ had reduced levels of cadherin-11 (CDH11), TIMP1, and heparin-binding EGF-like growth factor (HBEGF) before exercise. Whereas levels of endothelial cell-selective adhesion molecule (ESAM) were lower in ME/CFS^SD^ before and after exercise, levels of integrin α-6 (ITGA6) were elevated before exercise (**Fig. S6B**). Like cadherins, ESAM mediates cell-cell adhesion, but specifically between epithelial cells.^110,111^ TIMP1 released from activated astrocytes maintains the blood–brain barrier and protects neurons from excitotoxic injury.^112^ ITGA6, together with integrin β1 or β4, forms a receptor for laminin and mediates cell-ECM adhesion.^113,114^ ITGA6 elevation is consistent with enhanced ECM remodeling, consistent with TIMP1 depletion. HBEGF is a type I transmembrane protein from EGF family that is important in tissue repair and regeneration.^115^ HBEGF also protects against TNF-induced cell death in innate lymphoid tissues, reduces neuroinflammation in CNS, and increases glucose uptake and insulin sensitivity in muscle cells^116-119^.

In ME/CFS^LD^, levels of cadherin-1 (CDH1), bone morphogenetic protein 1 (BMP1), and WNT5A were elevated after exercise, while CCN4 levels were reduced at both time points (**Fig. S7B**). BMP1 is a metalloproteinase of the astacin family that plays an essential role in facilitating ECM turnover.^120^ WNT5A is a Wnt ligand that activates non-canonical Wnt signaling independent of β-catenin transcription.^121^ Activation of WNT5A-mediated non-canonical Wnt signaling induces adipose tissue inflammation, and serum levels of WNT5A correlate with levels of triglycerides and IL-6.^122,123^ WNT5A activation inhibits β-catenin accumulation and the Wnt canonical signaling pathway.^124,125^ CCN4 also regulates ECM homeostasis. It is a downstream target of Wnt1 and β-catenin in the canonical Wnt pathway.^126^ CCN4 can regulate its own expression by stimulating β-catenin activation.^127,128^ Pre- and post-exercise levels of CCN4 were reduced in ME/CFS^F^ (**Fig. S2B**). Baseline levels of CCN4 in ME/CFS^LD^ and the post-exercise levels in ME/CFS^F^ negatively correlated with MFI severity scores of mental fatigue and physical fatigue, respectively (**Fig. S7D, S2D**). CDH1 (E-cadherin) is one of the most critical molecules regulating cell-cell adhesion in epithelial tissues through interactions with β-catenin.^129^ MMPs directly cleave E-cadherin from the cell membrane, resulting in E-cadherin shedding into the bloodstream.^130^ Post-exercise levels of E-cadherin were also higher in ME/CFS and positively correlated with MFI reduced activity scores (**Fig. 3B**). Elevated levels of WNT5A and E-cadherin and reduced levels of CCN4 in ME/CFS^LD^ after exercise pointed to a disrupted Wnt signaling, indicating inhibition of canonical and activation of non-canonical Wnt signaling. Canonical Wnt actions are anti-adipogenic and anti-inflammatory, while non-canonical Wnt actions are pro-inflammatory and promotes adipogenesis and lipid accumulation.^131^ The observations of altered Wnt signaling pathways are consistent with the accumulation of triglycerides and IL-6 (**Fig. 4B**), and ECM disruption after exercise.

Like ME/CFS^F^, ME/CFS^OF^ had lower levels of CD93 and COMP (**Fig. S5B**). In concert, all subgroups of ME/CFS consistently showed significant reductions in molecules implicated in intercellular adhesion and immune cell trafficking, along with enhanced ECM breakdown and disrupted ECM homeostasis.

2.3.6. **Metabolic correlates of impaired resolution of inflammation in ME/CFS^YF^.**

Bioactive lipids are critically involved in resolving inflammation and restoring tissue integrity.^132^ Before exercise, ME/CFS^YF^, but not ME/CFS^OF^, had lower plasma levels of numerous bioactive lipids, including hydroxyoctadecadienoic acids (HODEs), hydroxyeicosatetraenoic acids (HETEs), hydroxyeicosapentaenoic acids (HEPEs), dihydroxyeicosatetraenoic acids (DIHETEs), epoxy eicosatrienoic acids (EpETrEs), and prostaglandins. Levels of ω3 polyunsaturated fatty acid (PUFA) docosahexaenoic acid (DHA) were also lower (**Fig. S4A**). HEPEs are derived from another ω3 PUFA eicosapentaenoic acid (EPA).^133^ The ω6 PUFAs arachidonic acid (AA) and linoleic acid (LA) are precursors to HODEs, HETEs, DiHETEs, EpETrEs, and prostaglandins.^134^ EPA and DHA exert their anti-inflammatory effects by competing with AA as substrates for COX-2 and 5-LOX enzymes.^135^ DHA gives rise to pro-resolving lipid mediators (SPMs), that facilitate the diminution and resolution of inflammation.^136^ Most AA-derived mediators promote immune cell activities.^137^ These baseline metabolomic findings suggest an overall deficiency in essential bioactive lipids, leading to inefficient resolution of inflammation and impaired immune responses. We previously reported elevated baseline levels of HEPEs in ME/CFS without age-stratification.^138^ The discrepancy may be due to the under-representation of ME/CFS^YF^ in the previous study, as only 26 of the 75 female patients (34.7%) were under 45 YoA. In ME/CFS^OF^, levels of prostaglandin reductase 1 (PTGR1) were higher before and after exercise (**Fig. S5B**). PTGR1 is a rate-limiting enzyme in AA metabolism that catalyzes the degradation of eicosanoids in resolution of inflammation.^139^ However, our metabolomic analysis did not discover differences in levels of eicosanoids between ME/CFS^OF^ and HC^OF^ (**Fig. S5A, Table S3E**).

2.3.7. **Proteomic correlates of abnormalities in adaptive immunity in ME/CFS^YF^ and** **ME/CFS^OF^.**

In ME/CFS^YF^, Th1 & Th2 pathways and pathways associated with signaling of IL-4 & IL-13, IL-12, IL-17, IL27, and IL-33 were altered both before and after exercise. IL-1 family signaling was altered after exercise. Th1 and Th2 pathways also exhibited an interaction effect (**Table S7D**). Baseline levels of TGF-β proproteins TGFB1 and TGFB2 were lower in ME/CFS^YF^ (**Fig. S4B**). TGF-β is mainly produced by immune cells and regulates cellular activities in the immune system.^140^ TGF-β renders a dual role as both a pro- and an anti-inflammatory mediator.^141^ In mice, TGF-β and IL-6 are required to produce IL-17 through Th-17 cells, a process crucial for the maintenance of gut homeostasis and mucosal barrier maintenance.^142,143^ Like TGFBs, baseline levels of IL6|IL6R were also reduced in ME/CFS^YF^ (**Fig. S4B**). By activating transcription factor Forkhead box P3 (FOXP3), TGF-β acts as an anti-inflammatory cytokine that induces Treg cell differentiation.^144^ FOXP3 is a master regulator that facilitates the development and suppressive function of Treg cells, even in the absence of TGF-β.^145^ Pro-inflammatory cytokines, like TNF-α and IL-6, degrade FOXP3.^146^ In contrast to TGFBs, baseline levels of FOXP3 were elevated in ME/CFS^YF^ (**Fig. S4B**). Activation of Treg cells enhances IL-10 signaling.^147^ We measured higher baseline levels of IL-10 receptor α (IL-10Rα) in ME/CFS^YF^(**Fig. S4B**). With engagement of IL-10Rα, Treg cells can more effectively suppress Th17 immunity.^148^ Reduced levels of TGFBs and IL6|IL6R, along with elevated levels of FOXP3 and IL-10Rα in ME/CFS^YF^ before exercise, may indicate enhancement of Treg cell activities and suppression of Th17 inflammatory response, leading to disrupted T cell homeostasis. In response to exercise, TGFB1 exhibited an increased interaction effect in ME/CFS^YF^, while FOXP3 showed a decreased interaction effect (**Table S6D**), potentially counteracting the baseline dysregulations. However, post-exercise levels of TGFB1 were still lower in ME/CFS^YF^ than in HC^YF^ (not significant).

After exercise, levels of IL-4, IL-11, and IL-23R were reduced in ME/CFS^YF^ (**Fig. S4B**). IL-4 is a regulatory cytokine that is critical in the type-2 immunity as it promotes Th2 differentiation but inhibits Th1 and Th17 differentiation.^149-151^ IL-4 binds to type I and type II receptors, both of which share IL-4 receptor subunit-α (IL-4Rα).^152^ The type I receptor is specific to IL-4 binding, while the type II IL-4R binds both IL-4 and IL-13.^153,154^ Soluble IL-4Rs antagonize IL-4 activity but do not block IL-13.^155^ In contrast to IL-4, IL-4Rα levels were elevated in ME/CFS^YF^ after exercise (**Fig. S4B**). No difference was noted in IL-13 levels (**Table S6D**). IL-11 is an IL-6 family regulatory cytokine that inhibits Th1 polarization but promotes Th2 polarization of naïve CD4+ T cells.**^156^** IL-11 can independently inhibit LPS-induced production of IL-1β, TNF-α, IFN-γ, and IL-12, thereby modulating effector function of macrophages.^157,158^ IL-23 is essential for Th17 cell function in inflammation.^159^ This process involves binding of IL-23 to IL-23R and subsequent activation of STAT3.^160^ Lower levels of IL-4, IL-11, and IL-23R in ME/CFS^YF^ suggest impairments in Th2 differentiation and Th17 effector function after exercise.

B cells produce antigen-specific antibodies that recognize and neutralize pathogens and are critical components in activating adaptive immunity.^161^ We and others have reported B cell expansion in subjects with ME/CFS.^162^ In ME/CFS^YF^, levels of joining chain (J-chain) were elevated both before and after exercise (**Fig. S4B**). J-chain is important for the formation and transport of polymeric IgA and IgM antibodies and for the regulation of immunity at mucosal linings.^163^ Previous studies on ME/CFS have reported elevated IgA and IgM antibody responses to commensal bacterial components.^34,164^ Lower pre- and post-exercise levels of FCGR1A in ME/CFS^F^ (see **Supplementary Materials 2.3.4** for innate immunity abnormalities in ME/CFS^F^) are consistent with impaired IgG-mediated antibody activities and functions.^165,166^ In the context of a compromised gut barrier (see **GI integrity in main text**), elevated J-chain levels and reduced FCGR1A levels in ME/CFS^YF^ reflect a pronounced non-specific polymeric IgA and IgM response across mucosal surfaces.

In ME/CFS^OF^, pathways involving IL-17A signaling in fibroblasts, IL-27 signaling, and IL-33 signaling were altered after exercise (**Fig. S5C, Table S7E**). Notably, post-exercise levels of C-C motif chemokine 1 (CCL1) and CCL8 were elevated (**Fig. S5B**). CCL1 and CCL8 both play critical roles in recruiting immune cells to sites of inflammation.^167,168^ These chemokines bind to the Th2 cell receptor CCR8 and promote the migration of Th2 cells and eosinophils.^169,170^ Consistently, higher post-exercise levels of IL-5, a Th2 cytokine, were observed in ME/CFS^OF^ (**Fig. S5B**). Serum levels of IL-5 were found to positively correlate with ME/CFS disease severity.^11^ These findings are consistent and indicate enhanced Th2 cell signaling in ME/CFS^OF^ after exercise.

2.3.8. **Subgroup-specific effects in mitochondrial dysfunction and lipid abnormalities.**
       2.3.8.1. **ME/CFS^YF^**

Before exercise, levels of cytochrome c-type heme lyase (HCCS), methylglutaconyl-CoA hydratase (AUH), and peptidyl-tRNA hydrolase 2 (PTRH2) were lower in ME/CFS^YF^ (**Fig. S4B**). HCCS is used for synthesis and maturation of cytochrome c, a critical component in electron transfer between complexes III and IV.^171^ AUH is a mitochondrial RNA-binding protein that regulates mitochondrial protein synthesis.^172^ AUH knockdown decreases the oxidase activity of cytochrome c and negatively affects mitochondrial respiration.^172^ PTRH2 is a mitochondrial protein that, by recycling peptidyl-tRNAs, maintains protein translation and synthesis.^173^ Depleted levels of HCCS, AUH, and PTRH2 suggest impairments within mitochondrial respiratory chain and oxidative phosphorylation (OXPHOS).

Phosphatidylcholines (PCs) are phospholipids that maintain the integrity of the inner mitochondrial membrane where OXPHOS occurs.^174^ Before exercise, ME/CFS^YF^ had lower levels of PC (32:1) and PC (33:1) than HC^YF^ (**Fig. S4A**). Baseline levels of phosphatidylcholine-sterol acyltransferase (LCAT) were elevated in ME/CFS^YF^ (**Fig. S4B**). LCAT is a central enzyme in the extracellular metabolism of plasma lipoproteins that converts cholesterol and PCs to cholesteryl esters and lyso-PCs.^175^ These findings confirm our previous reports of depletions in the plasma levels of PCs in ME/CFS at rest.^138,176^

After exercise, plasma levels of NAD-dependent protein deacetylase sirtuin-3 (SIRT3) and StAR-related lipid transfer protein 7 (STARD7) were elevated in ME/CFS^YF^ (**Fig. S4B**). SIRT3 is an exercise-induced protein in mitochondria that maintains redox balance through mitochondria biogenesis.^177^ Besides the deacetylation of mitochondrial proteins, it may also suppress mitochondrial protein synthesis.^178^ STARD7 delivers PCs from ER to mitochondria.^179^ Correspondingly, we observed decreased levels of PC (36:1) in ME/CFS^YF^ after exercise (**Fig. S4A**). PC reduction after exercise suggests more severe impairments in mitochondrial stability and energy production, concurrent with PEM.

In response to exercise, plasma levels of mitochondrial coenzyme Q6 monooxygenase (CoQ6) were increased in ME/CFS^YF^, but decreased in HC^YF^ (**Table S6D**). CoQ6 is required for the biosynthesis of CoQ10 and is an essential component of the mitochondrial electron transport chain (ETC).^180^ Increased levels of CoQ6 after exercise are consistent with increased oxidative stress. After exercise, levels of adenylate kinase isoenzyme 1 (AK1), an essential molecule regulating energy homeostasis, were decreased in ME/CFS^YF^ but increased in HC^YF^ (**Table S6D**). AK1 is found in cytosol of most tissues. It provides energy through catalysis of two ADP molecules to ATP and AMP and triggers cellular stress responses through the AMP signaling cascade.^181,182^ Similar to AK1, levels of phosphofructokinase (PFKM), pyruvate kinase (PKLR), ribose 5-phosphate isomerase A (RPIA), and acylphosphatase-1 (ACYP1) were decreased in ME/CFS^YF^ after exercise but increased in HC^YF^ (**Table S6D**). PFKM is a key regulatory enzyme in glycolysis that converts fructose-6-phosphate to fructose 1,6-*bis*phosphate.^183^ PKLR converts phosphoenolpyruvate to pyruvate and ATP.^184^ RPIA is a pentose phosphate pathway enzyme that provides an alternative route for hexose utilization.^185^ ACYP1 is a small cytosolic enzyme that catalyzes acyl phosphates and regulates metabolic pathways such as glycolysis and TCA cycle.^186,187^ In concert, these findings indicate impairments in the glycolytic and pentose phosphate pathways in ME/CFS^YF^ after exercise.^183,185,188,189^ In response to exercise, levels of lipoprotein lipase (LPL) were increased in ME/CFS^YF^ but decreased in HC^YF^ (**Table S6D**). Levels of patatin-like phospholipase domain-containing protein 2 (PNPLA2) were depleted in ME/CFS^YF^ after exercise (**Fig. S4B**). LPL is an enzyme that hydrolyzes triglycerides into free fatty acids and glycerol.^190^ PNPLA2, also known as adipose triglyceride lipase (ATGL), catalyzes lipolysis in an energy demanding environment.^191^ It is induced by exercise and regulates inflammatory responses.^192,193^ These proteomic findings may provide mechanistic insights into triglyceride accumulation.

2.3.8.2. **ME/CFS^OF^**

Before exercise, levels of the mitochondrial protein NDUFB8 were lower in ME/CFS^OF^ (**Fig. S5B**). NDUFB8 is an essential subunit of mitochondrial complex I and is involved in electron transport during OXPHOS.^194^ Levels of mitochondrial aspartate aminotransferase (GOT2) were higher in ME/CFS^OF^ before and after exercise (**Fig. S5B**; post-exercise elevation close to significance, **Table S6E**). GOT2 catalyzes the conversion of glutamate and OAA to aspartic acid and α-ketoglutarate.^195^ Elevated levels of GOT2 were consistent with increased levels of aspartic acid after exercise in ME/CFS^OF^ (**Table S3E**). Apolipoprotein E (APOE) is primarily noted for its role in lipid metabolism. It inhibits lipolysis, leading to triglyceride accumulation.^196,197^ Baseline levels of three major human isoforms of APOE (E2, E3, and E4) were higher in ME/CFS^OF^ (**Table S6E**).

Mitochondrial ribosomes are functionally specialized for the synthesis of mitochondrial membrane proteins, and hence, play critical roles in OXPHOS.^198^ MRPs are synthesized in the cytoplasm and are transported into mitochondria to be assembled into mitoribosome small (S) and large (L) subunits.^199^ Whereas baseline levels of MRPL21 were elevated in ME/CFS^OF^, levels of MRPL55 were lower in ME/CFS^SD^ before exercise and in ME/CFS^M^ after exercise (**Fig. S5B, S6B**, **S3B**). Post-exercise levels of MRPL33 were lower in ME/CFS^YF^ (**Fig. S4B**). Depletions in MRPs suggest impaired mitochondrial biogenesis.

2.3.9. **Metabolic correlates of neural abnormalities in ME/CFS subgroups.**
       2.3.9.1. **Dysregulated iron signaling in ME/CFS^F^.**

In ME/CFS^F^, plasma levels of ferritin (FTL ferritin light chain; FTL|FTH1 not differentiated ferritin light/heavy chain) were lower before and after exercise (**Fig. S2B**). Ferritin serves as the intracellular storage of iron, and low serum levels of ferritin reflect iron depletion.^200^ Iron deficiency may impair neurotransmission and functional plasticity of neurons.^201^ Although Yamamoto et al. reported higher serum levels of ferritin in female ME/CFS patients, low levels of ferritin have been historically associated with fatigue in both anemic and non-anemic individuals.^202,203^ In ME/CFS^F^, reduced baseline levels of ferritin correlated with MFI scores of physical fatigue and reduced motivation; post-exercise levels correlated with MFI scores of reduced motivation (**Fig. S2D**).

2.3.9.2. **Dysregulated NAD+ signaling in ME/CFS^M^.**

In ME/CFS^M^, levels of nicotinamide riboside kinase (NMRK1) were reduced before exercise (**Fig. S3B**). NMRK1 is the rate-limiting enzyme that catalyzes the conversion of nicotinamide riboside (NR) to NAD+, downstream of the kynurenine (KYN) pathway. ^204-206^ NAD+ is indispensable for energy metabolism, DNA damage repair, as well as cell signaling and survival.^207^ Reduced NAD+ levels have been reported in ME/CFS; interventions to raise intracellular NAD+ levels have been proposed to improve neurological function, promote energy production, and relieve fatigue in ME/CFS.^208^ In addition, ME/CFS^M^ had reduced baseline levels of tyrosine kinase receptor ERBB3. Levels of neuregulin-3 (NRG3) were reduced both before and after exercise. Levels of tropomyosin alpha-1 chain (TPM1) were elevated after exercise (**Fig. S3B**). NRGs are implicated in the pathogenesis of neurodegenerative diseases.^209,210^ NRG3 is a neuronal-enriched neurotrophin that plays a role in regulating the activation of its receptor ERBB4 in the central nervous system (CNS).^211^ Ablation of NRG3 leads to reduced formation of excitatory synapse on hippocampal interneurons.^212^ In ME/CFS^M^, post-exercise levels of NRG3 inversely correlated with MFI scores of general and mental fatigue (**Fig. S3D**). ERBB receptors, when activated, initiate intracellular signaling cascades, including PI3K/AKT and MAPK/ERK pathways, contributing to inflammation and immune infiltration.^213,214^ ERBB3 binds NRG1, a protein structurally similar to NRG3, and maintains blood brain barrier.^215^ TPM1 is an actin-dependent muscle protein that can regulate inflammation downstream of TREM2.^216^ TPM1 accumulation has been associated with neuronal remodeling and inflammation in mouse models.^217^ Taken together, these findings in ME/CFS^M^ suggest impaired intracellular signaling that may contribute to disrupted synapse formation, altered neuronal connectivity, and increased susceptibility to neuroinflammation.

2.3.10. **Peroxisomal dysfunction in ME/CFS subgroups.**

Plasmalogens, constituting up to 20% of the total phospholipid mass, are synthesized in peroxisomes and ER, and play key roles in signal transduction and mitigating ROS.^218^ We have previously reported reduced plasma levels of plasmalogens in ME/CFS at rest.^138^ We confirmed here that, before exercise, levels of two structural isomers of PC (p-42:5)/PC (o-42:6) and very-long-chain PC ethers (PC-ether-vlc) were lower in ME/CFS^M^ (**Fig. S3A**). Proteomic analysis detected increased levels of phytanoyl-CoA dioxygenase (PHYH) in ME/CFS following exercise (**Table S6A**). PHYH is an important enzyme for peroxisomal α-oxidation,^219^ and increased PHYH levels are consistent with higher propensity of α-oxidation. Pipecolic acid is a γ-aminobutyric acid (GABA) receptor agonist metabolized in peroxisomes.^220^ Higher levels of pipecolic acid indicate peroxisomal dysfunction.^221^ Whereas pipecolic acid levels were increased in ME/CFS^F^ after exercise, levels in HC^F^ were decreased (**Table S3B**). In ME/CFS^SD^, levels of metabolites in PC-ether, PC-ether-vlc, and PUFA phosphatidylethanolamine (PE) and PC ether (PE-ether-PUFA, PC-ether-PUFA) were either elevated or increased after exercise (**Fig. S6A**). Donovan et al. reported higher plasma levels of ether lipids in obese individuals that correlated with higher oxidative stress^222^. After exercise, levels of peroxisomal oxidase (PIPOX) were also increased in ME/CFS^SD^ (**Fig. S6B**). PIPOX catalyzes the oxidation of sarcosine into glycine and formaldehyde, wherein H_2_O_2_ is produced.^223^ Increased levels of ether lipids and PIPOX suggest a prolonged oxidative state in ME/CFS^SD^ following exercise.

In ME/CFS^YF^, levels of catalase (CAT) were decreased after exercise but increased in HC^YF^ (**Table S6D**). Post-exercise levels of cytosolic superoxide dismutase (SOD1) were lower (**Fig. S4B**). CAT is a peroxisomal antioxidant enzyme that neutralizes ROS.^224^ Another antioxidant, SOD1, regulates the transcription of antioxidant pathways.^225^ NADPH-dependent cytochrome P450 reductase (POR), located on ER membrane, regulates xenobiotic metabolism and lipid homeostasis while generating ROS as a "leaky enzyme" through NADPH oxidation.^226^ In ME/CFS^YF^ after exercise, levels of POR were elevated compared to HC^YF^ (**Fig. S4B**).

2.3.11. **Abnormalities in methionine cycle and glutathione (GSH) biosynthesis.**

In metabolomic analysis, plasma levels of methionine were increased following exercise in HC, but not in ME/CFS (**Fig. 2D**). Methionine is an essential amino acid that, through the formation of S-adenosylmethionine (SAM), plays a key role in the one-carbon metabolism.^227^ Proteomic analysis revealed post-exercise abnormalities in the methionine cycle enzymes. After exercise, levels of S-adenosylmethionine synthase isoform type-1 (MAT1A) and adenosylhomocysteinase (AHCY) were both higher in ME/CFS^OF^ (**Fig. S5B**). MAT1A is part of methionine adenosyltransferase (MAT) that catalyzes the biosynthesis of SAM from methionine.^228^ SAM donates its methyl group and generates S-denosylhomocysteinee (SAH) which is then hydrolyzed into homocysteine and adenosine through AHCY.^229^ Elevated post-exercise levels of MAT1A and AHCY indicate increased metabolism from methionine to homocysteine in ME/CFS^OF^, a finding in line with increased levels of methionine in HC, but not in ME/CFS. Levels of serine were higher after exercise in ME/CFS (**Fig. 2B**). Serine is produced through the actions of phosphoglycerate dehydrogenase (PGDH), phosphoserine aminotransferase (PSAT1), and phosphoserine phosphatase (PSPH).^230,231^ We found higher post-exercise levels of PSAT1 in ME/CFS^SD^ (**Fig. S6B**). PSPH levels were elevated in ME/CFS after exercise and correlated with MFI physical fatigue scores (**Fig. 2F, 3B**). Increased levels of PSAT1 and PSPH are consistent with higher levels of serine in ME/CFS after exercise. Enhanced metabolism from methionine to homocysteine and elevated levels of serine may point to an activation of the transsulfuration pathway, wherein homocysteine and serine serve as two main precursors of cysteine.^232^ Correspondingly, levels of cystathionine beta-synthase (CBS), the rate-limiting enzyme of the transsulfuration pathway,^232^ were elevated in ME/CFS^LD^ both before and after exercise (**Fig. S7B**).

Cysteine can be metabolized into GSH, an antioxidant that regulates cellular metabolism and tissue repair.^233^ Lower plasma levels of GSH were reported in ME/CFS at rest.^234^ The enzymic action of GSH synthesis is carried out by glutamate cysteine ligase (GCL).^235^ GCL consists of catalytic (GCLC) and modifier (GCLM) subunits, while GCLM alone can increase the catalytic efficiency of cellular GCL activity.^236,237^ Whereas GCLM levels were decreased after exercise in ME/CFS^YF^, they were increased HC^YF^ (interaction, BF=4.212). GSH conjugation via glutathione-S-transferase (GST) is a detoxification pathway for electrophilic xenobiotics.^238^ GSTs are characterized into various classes: *alpha* (GSTA), *mu* (GSTM), *pi* (GSTP), *theta* (GSTT), and *zeta* (GSTZ).^239^ We found lower plasma levels of GSTZ1 in ME/CFS^YF^ before and after exercise (**Fig. S4B**). GSTZ1 serves as a fundamental enzyme in tyrosine metabolism.^240,241^ *Gstz1*^-/-^ knockout mice have reduced GSH concentrations and increased activities of GSTA and GSTM.^242^ Consistent with this transgenic model, pre- and post-exercise levels of GSTA1 were elevated in ME/CFS^F^ (**Fig. S2B**); levels of GSTM1 were elevated in ME/CFS^YF^ before exercise and in ME/CFS^SD^ after exercise (**Fig. S4B, S6B**). GSTZ1 deficiency leading to glutathione depletion may induce oxidative stress and activates the adaptive antioxidant response pathways in ME/CFS. Antioxidant activities of GSH involve glutathione peroxidase (GPX).^243^ Levels of GPX5 were higher in ME/CFS both before and after exercise (**Fig. 2E, 2F**), accordant with increased oxidative stress. These findings reflect disruptions of methionine to GSH biosynthesis pathways that might result in inefficient antioxidation.

2.3.12. **Proteomic evidence of abnormalities in** **UPS in ME/CFS^YF^ after exercise.**

Ubiquitin-proteasome system (UPS) maintains protein quality control and cellular homeostasis by tagging damaged and misfolded proteins for degradation.^244^ Plasma proteomic analysis indicated UPS disruption in ME/CFS^YF^ after exercise (**Table S7D**). In response to exercise, levels of polyubiquitin-B (UBB) and -C (UBC), ubiquitin-conjugating enzymes UBE2E3, UBE2L6, and UBE2V1, 20S proteasome subunit PSMA7, and 26S proteasome subunits PSMC3 and PSMD9 all exhibited a decreased interaction effect, where the levels were decreased in ME/CFS^YF^ but increased in HC^YF^ (**Table S6D**). In the UPS, ubiquitin selectively tags proteins for degradation by the 26S proteasome.^245,246^ UBB is essential for the survival of neurons, and mice lacking UBB developed a progressive neurodegenerative disorder.^247^ UBC plays a unique role in maintaining cellular ubiquitin levels.^248^ Ubiquitin-conjugating enzymes (E2s) attach ubiquitin to cellular proteins.^249^ UBE2E3 is restricted to generating monoubiquitylated products and is involved in antioxidant pathways.^250,251^ Induced by IFN-γ, UBE2L6 promotes polyubiquitylation through its conjugation activity in response to IFNs.^252^ The E2 variant UBE2V1 acts as a cofactor for the E2 enzyme UBE2N, and their interaction is required for the activation of NF-κB and MAPK signaling pathways.^253,254^ It promotes protein aggregate formation and mediates FGF signaling.^255,256^ Besides modulating protein degradation, 20S proteasome subunit PSMA7 can also promote NF-κB and MAPK activation.^257,258^ Both PSMC3 and PSMD9 are subunits of the 19S regulatory particle, which recognizes and binds polyubiquitinated proteins.^259^ PSMC3 is an ATP-dependent subunit that is critically involved in CNS development and cognitive function through type I interferon signaling.^260^ The non-ATPase subunit PSMD9 serves as one of the chaperones that mediates the assembly of the 26S proteasome complex.^261^ In concert, these findings point to an overall downregulation of the UPS in ME/CFS^YF^ in response to exercise, leading to disrupted cellular homeostasis and impaired stress response. HLA-G is a non-classical MHCI protein. Its expression is induced in EBV-transformed B cells under conditions of nutrient deficiency, hypoxia, or both, suggesting that HLA-G is stress-responsive.^262,263^ Consistent with elevated cellular stress due to impaired UPS, HLA-G levels had an increased interaction effect in ME/CFS^YF^ after exercise (**Table S6D**).

2.3.12.1. **MAPK activation in ME/CFS^YF^ and ME/CFS^OF^ after exercise.**

In ME/CFS^YF^, levels of p38 MAPK-activating factors MAP2K6 and MAP3K3 were elevated both before and after exercise (**Fig. S4B**). MAP3K3 promotes the phosphorylation of MAP2K6 which activates the p38 MAPK signal transduction pathway.^264^ A prolonged activation of p38 MAPK pathway reflects cellular and oxidative stress and induces inflammatory responses in ME/CFS^YF^ after exercise.^265,266^ In ME/CFS^OF^, plasma levels of MAPK8 were elevated before and after exercise (**Fig. S5B**). MAPK8 is activated by cellular stress and regulates immune responses.^267^ As a key priming event, it phosphorylates NLRP3 directly and activates NLRP3 inflammasome, leading to productions of IL-1β and caspase-1.^268^ MAPK activations in ME/CFS^YF^ and ME/CFS^OF^ provide additional support for differential inflammatory mediators within females with ME/CFS.

2.3.12.2. **Abnormalities in the metabolism of gut-derived phenylacetylglutamine.**

Polyphenols are natural compounds that are metabolized by gut microbial communities.^269^ The metabolism of polyphenols yields bioactive molecules, most of which have beneficial roles in regulating oxidative stress and inflammation.^270^ One phenolic metabolite worth noting is phenylacetylglutamine (PAGln). PAGln is produced by gut bacteria in the transformation of phenylalanine into phenylacetic acid, followed by conjugation of glutamine in the liver.^271^ PAGln acts as an activator for platelets through adrenergic receptors and increase risk to thrombosis.^272^ PAGln also serves as an alternative to urea as a vehicle for nitrogen waste excretion.^273^ Whereas plasma levels of PAGln were increased in ME/CFS^YF^ after exercise, they were decreased in HC^YF^ (**Table S3D**). Levels of glutamine were increased in ME/CFS^OF^ after exercise while decreased in HC^OF^ (**Table S3E**). In patients with hyperammonemia, accumulation of nitrogen waste in the blood leads to increased levels of glutamine.^274^ With a disrupted urea cycle, elevated levels of PAGln and glutamine imply further post-exercise abnormalities in platelet activation and removal of nitrogen waste in ME/CFS^F^.

2.3.13. **Sex-specific hormonal imbalance.**

Although higher serum levels of insulin have been described in ME/CFS,^275^ the influence of exercise on insulin levels has not been reported. In the proteomic analysis, we confirmed elevated levels of insulin before exercise in ME/CFS^M^ (**Fig. S3B**). The baseline elevation correlated with MFI mental fatigue scores (**Fig. S3D**). No baseline differences were observed between ME/CFS^F^ and HC^F^. After exercise, insulin levels decreased in HC^YF^, but not in ME/CFS^YF^ (**Fig. S4B**). Levels of insulin-degrading enzyme (IDE) were decreased in ME/CFS^YF^ while increased in HC^YF^ (interaction, BF=3.482). Decreased insulin levels in the healthy population suggest enhanced glucose utilization for energy production following exercise. However, this regulatory mechanism may be impaired in ME/CFS^YF^. Estrogens also enhance insulin sensitivity.^276^ Accordingly, we observed decreased levels of insulin after exercise in HC^YF^, but not in their older counterparts who had lower plasma levels of E2 (**Table 2**). IDE serves as a major proteolytic enzyme for the clearance of both extracellular and intracellular amyloid beta (Aβ) peptides.^277^ IDE deficiency has been implicated as a trigger in AD**.**^278^ Fibrinaloid microclots comprised of amyloid have been reported in blood of ME/CFS patients.^279^ Furthermore, Baraniuk et al. have detected amyloidogenic proteins in the cerebrospinal fluid (CSF) of patients with ME/CFS, Gulf War Illness (GWI) and fibromyalgia (FM).^280^

Levels of insulin, glucagon, and other peptide hormones are modulated by enzymes that convert cognate prohormones to active hormones. Neuroendocrine convertase 1 and 2 (PCSK1 and PCSK2) convert proinsulin to insulin and other neuroactive peptides.^281^ Consistent with higher baseline levels of insulin, levels of PCSK2 were elevated before and after exercise in ME/CFS^M^ (**Fig. S3B**). In contrast, levels of PCSK1 were depleted in ME/CFS^YF^ before exercise (**Fig. S4B**). These findings may reflect differences in the conversion from prohormones to active hormones between ME/CFS^F^ and ME/CFS^M^. ME/CFS^LD^ had lower levels of iron-regulating hormone hepcidin (HAMP) before exercise (**Fig. S7B**). Kavyani et al. also found reduced plasma levels of HAMP in ME/CFS.^282^ Its reduction reflects systemic lower amount of iron and may imply ongoing hypoxia.^282,283^

The hypothalamic-pituitary-adrenal (HPA) axis is the primary stress response system.^284^ Previous studies have reported lower HPA activity and decreased levels of cortisone and cortisol in ME/CFS.^285-287^ Cortisol is the active form of the glucocorticoid hormone derived from the enzymatic conversion of cortisone through 11β-hydroxysteroid dehydrogenase 1(HSD11B1).^288^ Cortisol can also be converted to cortisone by 11β-hydroxysteroid dehydrogenase 2 (HSD11B2).^288^ In ME/CFS^M^, levels of HSD11B1 were elevated after exercise (**Fig. S3B**). Whereas levels of cortisone were depleted in ME/CFS^F^ after exercise (**Fig. S2A**, levels of cortisol and HSD11B1 were similar between ME/CFS^F^ and HC^F^ (**Table S3B**). Levels of progesterone, a gonadal steroid, were lower after exercise in ME/CFS^YF^ (**Fig. S4A**). Under stress (exercise), cortisol and progesterone play important roles in regulating energy homeostasis and memory functions.^289,290^

Immune-mediated loss of thyroid function has been reported in ME/CFS.^291^ Ruiz-Núñez et al. found lower serum triiodothyronine (T3) and thyroxine (T4) levels and a higher prevalence of “low T3 syndrome” in ME/CFS patients.^292^ Thyroglobulin (TG) is the protein precursor for the synthesis of the peripheral T3 and T4.^293^ In our proteomic analysis, levels of TG were lower before exercise in ME/CFS^M^ (**Fig. S3B**). Thyroid-stimulating hormone (TSH) can stimulate the production of TG.^294^ Glycoprotein hormone α chain (CGA) and thyrotropin subunit β (TSHB) provide the structural support for all glycoprotein hormones, including TSH, follicle-stimulating hormones (FSH), and luteinizing hormone (LH). Our proteomic analysis also revealed decreased plasma levels of CGA|TSHB in ME/CFS after exercise (**Fig. S3B**).

2.4. **Analyte-analyte correlations in ME/CFS and HC.**

In ME/CFS before exercise, 1-methygalactose, linalool, HTRA1, together with di- and triglycerides were correlated within the same module with TAG (56:5) C as the center, suggesting their interactive roles in inflammation; indole-3-lactate (ILA) and glucuronic acid were clustered together reflecting abnormalities in the gut microbiome; cell surface receptors EPHA4, KDR, and SELL, all critically involved in cell-cell interactions, were closely inter-correlated. In HC, ILA correlated with organic acids; SELL coordinated a module with 12,13-diHOME, HTRA1, and BATF3 were correlated in the same module, indicating their roles in the regulation of immune/metabolic processes (**Fig. 3E**). After exercise, whereas DAG (36:3) and TAG (56:6) were clustered together in ME/CFS, they also correlated with serine and PSPH (enzyme for serine biosynthesis) in HC. Differences in the inter-analyte networks between ME/CFS and HC after exercise, highlighted altered metabolic and immune regulation in ME/CFS, with disrupted lipid-inflammatory interactions and impaired serine biosynthesis (**Supplementary Materials 2.3.11** for abnormalities in methionine cycle and GSH biosynthesis).

2.5. **External Validation for Metabolomics.**

External validation^30^ for plasma metabolomic analysis was conducted on the biological pathway level using MetaboAnalyst 6.0,^295^ identifying significant pathways (p<0.1) comparing ME/CFS vs. HC before and 24 hours after exercise. A total of 566 metabolites from our study and 562 metabolites from the external study^30^ were annotated with Human Metabolome Database (HMDB) IDs. MetaboAnalyst identified same pathways in both studies, including those involved in amino acid metabolism, fatty acid degradation and synthesis, lipid metabolism, and xenobiotic detoxification (**Table S10**). When comparing ME/CFS^YF^ vs. HC^YF^, 3 (out of 5) pre-exercise pathways and 3 (out of 4) post-exercise pathways that were significant in our study were identified in the external validation study (**Table S10D**). Aside from the disparities between the two metabolomic assays (only 220 matched metabolites), the validation analysis was also confounded by differences in subject characteristics. While we collected similar sample numbers from ME/CFS^YF^ (n=18) and ME/CFS^OF^ (n=19), Germain et al. collected more samples from ME/CFS^OF^ (n=26) than from ME/CFS^YF^ (n=19).^30^

**Supplementary Figure Legends**

**Figure S1. No discriminatory inflammatory responses to LPS and poly I:C.**

**(A).** **ME/CFS vs HC:** Box plots showing concentrations of cytokines (pg/µl) in blood exposed to LPS (left) and poly I:C (right) before and 24 hours after exercise. Comparisons employed linear mixed-effect models (LMMs) paired with Bayesian analyses, adjusted for age, sex, race/ethnicity, geography, body mass index (BMI), and self-reported irritable bowel syndrome (sr-IBS). The full TruCulture cytokine results are reported in **Table S8.**

**(B). Sex- and age- stratified analysis:** Box plots showing sex-and age-stratified concentrations of cytokines (pg/µl) in blood exposed to LPS (up) and poly I:C (down) before and 24 hours after exercise. Comparisons employed linear mixed-effect models (LMMs) paired with Bayesian analysis in females and males, separately. The full TruCulture cytokine results are reported in **Table S8**.

**Figure S2. Multi-omics analyses comparing ME/CFS^F^ vs. HC^F^.**

**(A).** Scatter plots showing BayesFactors (BFs) associated with differences in plasma levels of metabolomic analytes between ME/CFS^F^ vs. HC^F^ before and 24 hours after exercise, as well as changes in plasma levels of metabolites from before to 24 hours after exercise in ME/CFS^F^ and in HC^F^. The full metabolomic results, comparing ME/CFS^F^ and HC^F^, are reported in **Table S3B**.

**(B).** Volcano plots showing BayesFactors (BFs) associated with differences in plasma levels of proteomic analytes between ME/CFS^F^ vs. HC^F^ before and 24 hours after exercise, as well as changes in plasma levels of proteomic analytes from before to 24 hours after exercise in ME/CFS^F^. No proteomic analytes showed significant changes in the levels from before to after exercise in HC^F^. The full proteomic results, comparing ME/CFS^F^ and HC^F^, are reported in **Table S6B**.

**(C).** Ingenuity pathway analyses (IPA) combining the metabolomic and proteomic analytes with significant differences between ME/CFS^F^ and HC^F^ before and 24 hours after exercise. The full IPA results , comparing ME/CFS^F^ and HC^F^, are reported in **Table S7B**.

**(D).** Alluvial plots showing the regularized canonical correlations between MFI symptom severity scales and plasma levels of metabolomic and proteomic analytes, which showed significant difference between ME/CFS^F^ and HC^F^ after exercise, in ME/CFS^F^.

**Figure S3. Multi-omics analyses comparing ME/CFS^M^ vs. HC^M^.**

**(A).** Scatter plots showing BayesFactors (BFs) associated with differences in plasma levels of metabolomic analytes between ME/CFS^M^ vs. HC^M^ before and 24 hours after exercise, as well as changes in plasma levels of metabolites from before to 24 hours after exercise in ME/CFS^M^ and in HC^M^. The full metabolomic results, comparing ME/CFS^M^ and HC^M^, are reported in **Table S3C**.

**(B).** Volcano plots showing BayesFactors (BFs) associated with differences in plasma levels of proteomic analytes between ME/CFS^M^ vs. HC^M^ before and 24 hours after exercise, as well as changes in plasma levels of proteomic analytes from before to 24 hours after exercise in ME/CFS^M^ and in HC^M^. The full proteomic results, comparing ME/CFS^M^ and HC^M^, are reported in **Table S6C**.

**(C).** Ingenuity pathway analyses (IPA) combining the metabolomic and proteomic analytes with significant differences between ME/CFS^M^ and HC^M^ before and 24 hours after exercise. The full IPA results, comparing ME/CFS^M^ and HC^M^, are reported in **Table S7C**.

**(D).** Alluvial plots showing the regularized canonical correlations between MFI symptom severity scales and plasma levels of metabolomic and proteomic analytes, which showed significant difference between ME/CFS^M^ and HC^M^ after exercise, in ME/CFS^M^.

**Figure S4. Multi-omics analyses comparing ME/CFS^YF^ vs. HC^YF^.**

**(A).** Scatter plots showing BayesFactors (BFs) associated with differences in plasma levels of metabolomic analytes between ME/CFS^YF^ vs. HC^YF^ before and 24 hours after exercise, as well as changes in plasma levels of metabolites from before to 24 hours after exercise in ME/CFS^YF^ and in HC^YF^. The full metabolomic results, comparing ME/CFS^YF^ and HC^YF^, are reported in **Table S3D**.

**(B).** Volcano plots showing BayesFactors (BFs) associated with differences in plasma levels of proteomic analytes between ME/CFS^YF^ vs. HC^YF^ before and 24 hours after exercise, as well as changes in plasma levels of proteomic analytes from before to 24 hours after exercise in ME/CFS^YF^ and in HC^YF^. The full proteomic results, comparing ME/CFS^YF^ and HC^YF^, are reported in **Table S6D**.

**(C).** Ingenuity pathway analyses (IPA) combining the metabolomic and proteomic analytes with significant differences between ME/CFS^YF^ and HC^YF^ before and 24 hours after exercise. The full IPA results, comparing ME/CFS^YF^ and HC^YF^, are reported in **Table S7D**.

**(D).** Alluvial plots showing the regularized canonical correlations between MFI symptom severity scales and plasma levels of metabolomic and proteomic analytes, which showed significant difference between ME/CFS^YF^ and HC^YF^ after exercise, in ME/CFS^YF^.

**Figure S5. Multi-omics analyses comparing ME/CFS^OF^ vs. HC^OF^.**

**(A).** Scatter plots showing BayesFactors (BFs) associated with differences in plasma levels of metabolomic analytes between ME/CFS^OF^ vs. HC^OF^ before and 24 hours after exercise, as well as changes in plasma levels of metabolites from before to 24 hours after exercise in ME/CFS^OF^ and in HC^OF^. The full metabolomic results, comparing ME/CFS^OF^ and HC^OF^, are reported in **Table S3E**.

**(B).** Volcano plots showing BayesFactors (BFs) associated with differences in plasma levels of proteomic analytes between ME/CFS^OF^ vs. HC^OF^ before and 24 hours after exercise, as well as changes in plasma levels of proteomic analytes from before to 24 hours after exercise in ME/CFS^OF^ and in HC^OF^. The full proteomic results, comparing ME/CFS^OF^ and HC^OF^, are reported in **Table S6E**.

**(C).** Ingenuity pathway analyses (IPA) combining the metabolomic and proteomic analytes with significant differences between ME/CFS^OF^ and HC^OF^ before and 24 hours after exercise. The full IPA results, comparing ME/CFS^OF^ and HC^OF^, are reported in **Table S7E**.

**(C).** Alluvial plots showing the regularized canonical correlations between MFI symptom severity scales and plasma levels of metabolomic and proteomic analytes, which showed significant difference between ME/CFS^OF^ and HC^OF^ after exercise, in ME/CFS^OF^.

**Figure S6. Multi-omics analyses comparing ME/CFS^SD^ vs. HC.**

**(A).** Scatter plots showing BayesFactors (BFs) associated with differences in plasma levels of metabolomic analytes between ME/CFS^SD^ vs. HC before and 24 hours after exercise, as well as changes in plasma levels of metabolites from before to 24 hours after exercise in ME/CFS^SD^. The full metabolomic results, comparing ME/CFS^SD^ and HC, are reported in **Table S3F**.

**(B).** Volcano plots showing BayesFactors (BFs) associated with differences in plasma levels of proteomic analytes between ME/CFS^SD^ vs. HC before and 24 hours after exercise, as well as changes in plasma levels of proteomic analytes from before to 24 hours after exercise in ME/CFS^SD^. The full proteomic results, comparing ME/CFS^SD^ and HC, are reported in **Table S6F**.

**(C).** Ingenuity pathway analyses (IPA) combining the metabolomic and proteomic analytes with significant differences between ME/CFS^SD^ and HC before and 24 hours after exercise. The full IPA results, comparing ME/CFS^SD^ and HC, are reported in **Table S7F**.

**(D).** Alluvial plots showing the regularized canonical correlations between MFI symptom severity scales and plasma levels of metabolomic and proteomic analytes, which showed significant difference between ME/CFS^SD^ and HC after exercise, in ME/CFS^SD^.

**Figure S7. Multi-omics analyses comparing ME/CFS^LD^ vs. HC.**

**(A).** Scatter plots showing BayesFactors (BFs) associated with differences in plasma levels of metabolomic analytes between ME/CFS^LD^ vs. HC before and 24 hours after exercise, as well as changes in plasma levels of metabolites from before to 24 hours after exercise in ME/CFS^LD^. The full metabolomic results, comparing ME/CFS^LD^ and HC, are reported in **Table S3G**.

**(B).** Volcano plots showing BayesFactors (BFs) associated with differences in plasma levels of proteomic analytes between ME/CFS^LD^ vs. HC before and 24 hours after exercise, as well as changes in plasma levels of proteomic analytes from before to 24 hours after exercise in ME/CFS^LD^. The full proteomic results, comparing ME/CFS^LD^ and HC, are reported in **Table S6G**.

**(C).** Ingenuity pathway analyses (IPA) combining the metabolomic and proteomic analytes with significant differences between ME/CFS^LD^ and HC before and 24 hours after exercise. The full IPA results, comparing ME/CFS^LD^ and HC, are reported in **Table S7G**.

**(D).** Alluvial plots showing the regularized canonical correlations between MFI symptom severity scales and plasma levels of metabolomic and proteomic analytes, which showed significant difference between ME/CFS^LD^ and HC after exercise, in ME/CFS^LD^.
