## Supplementary figures and images for "Heightened innate immunity may trigger chronic inflammation, fatigue and post-exertional malaise in ME/CFS"

### Supplement figures

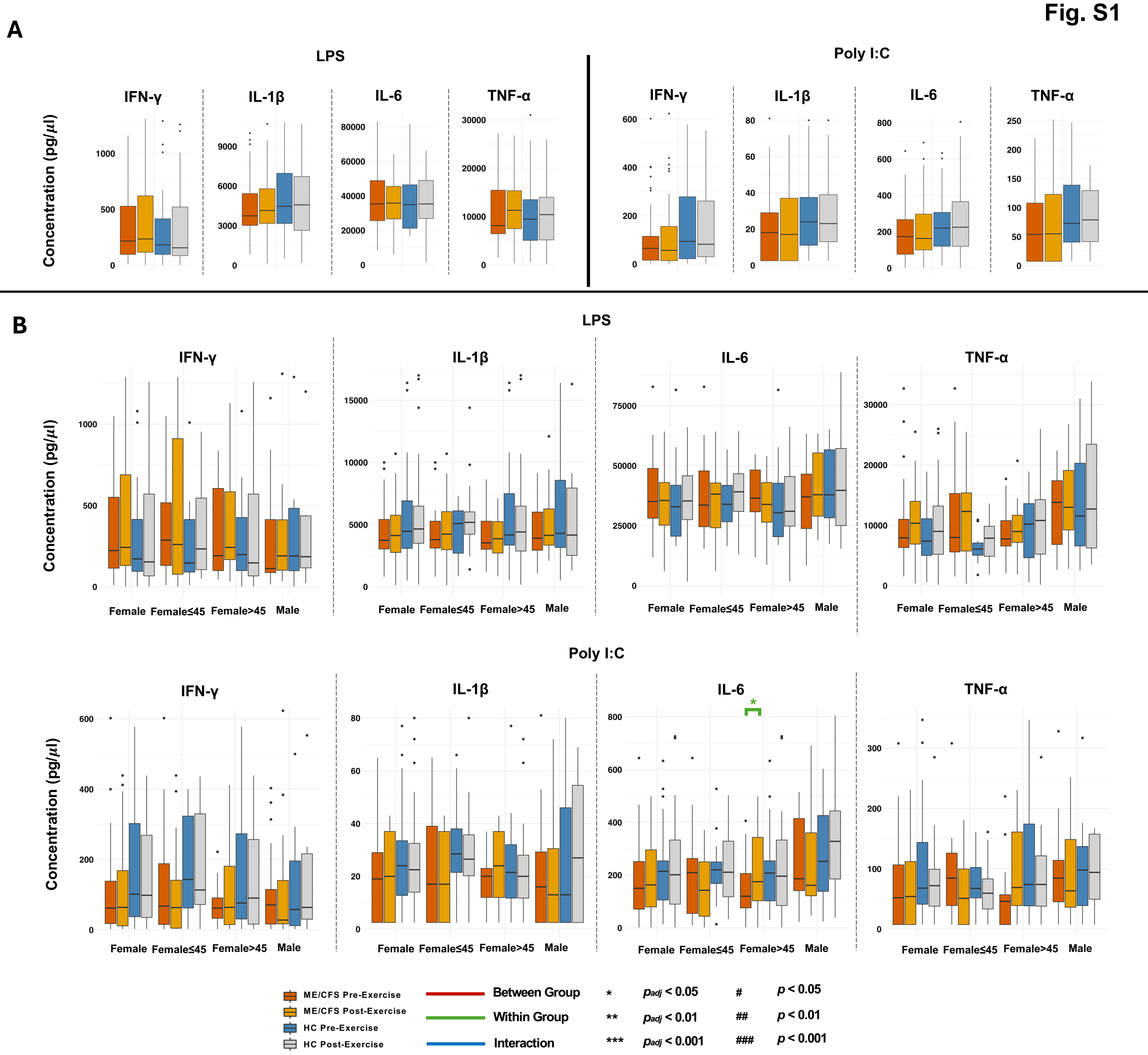

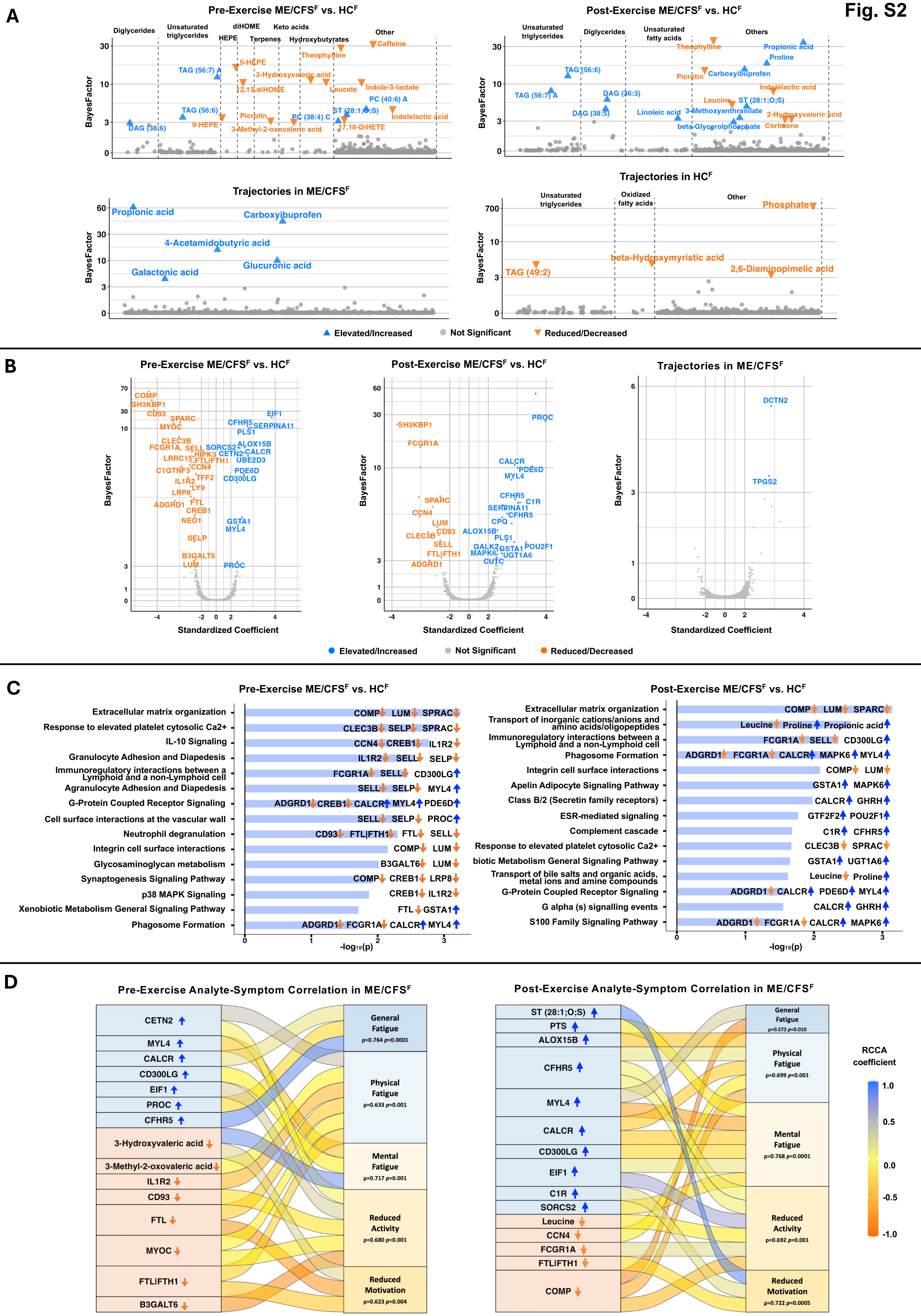

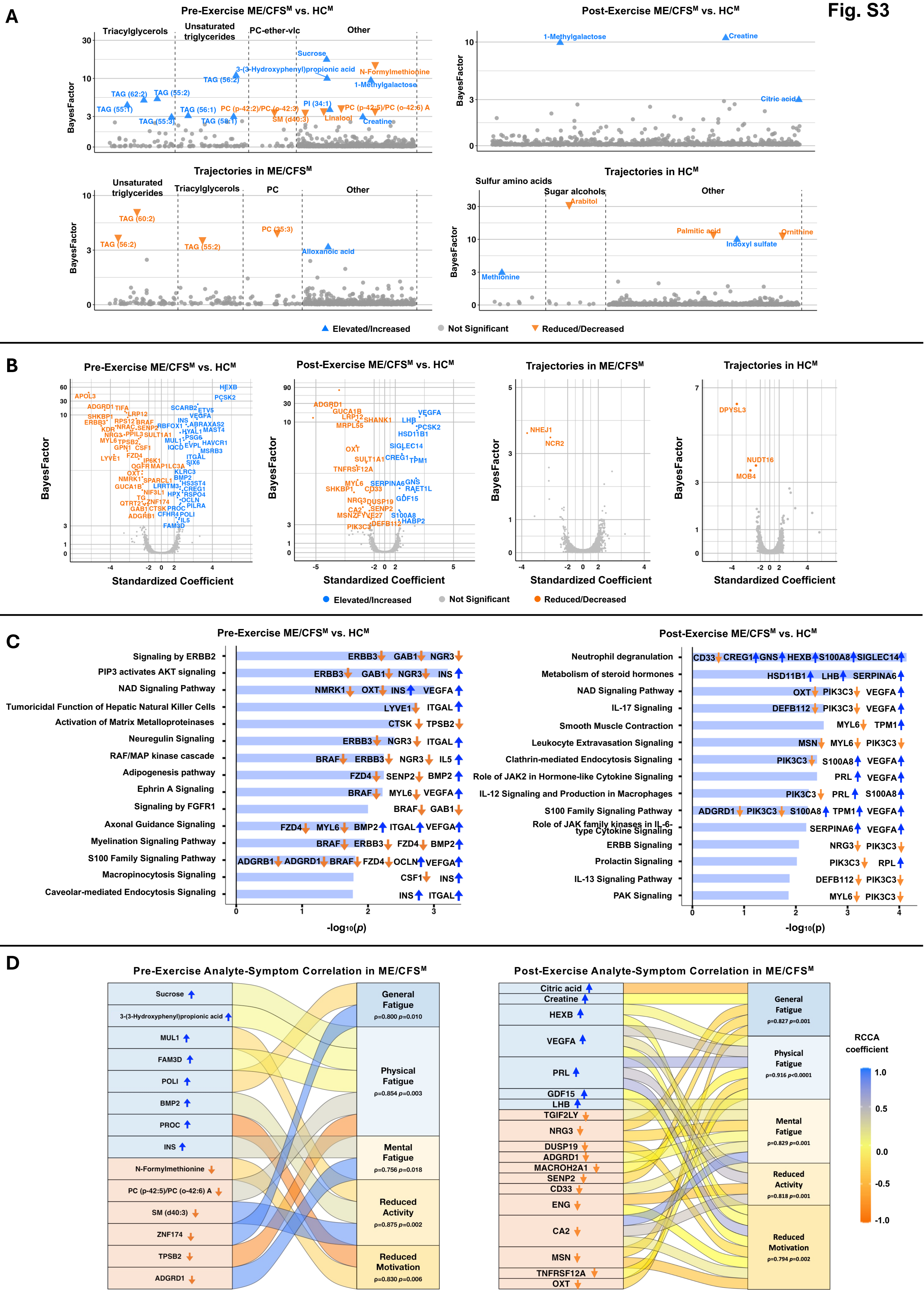

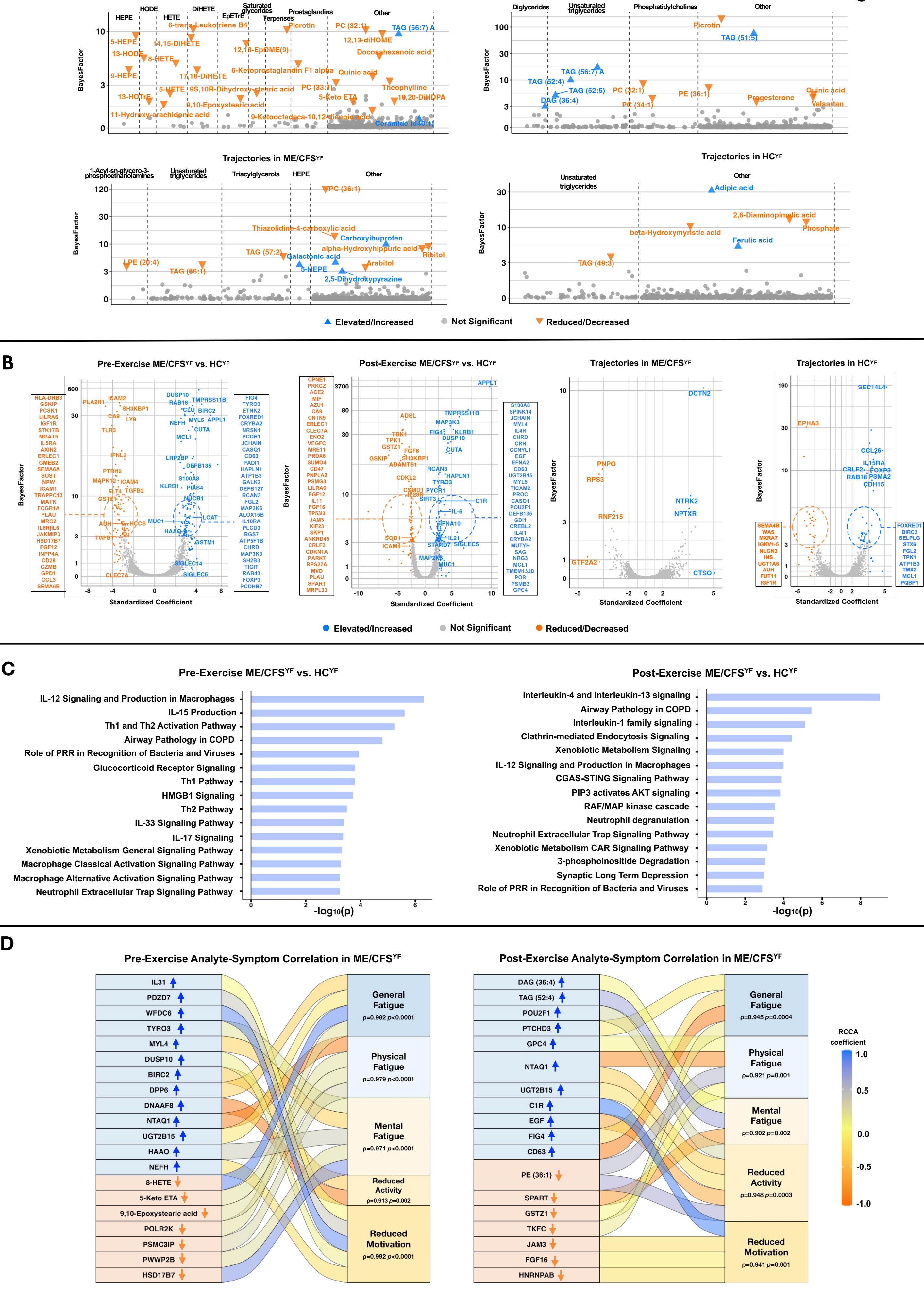

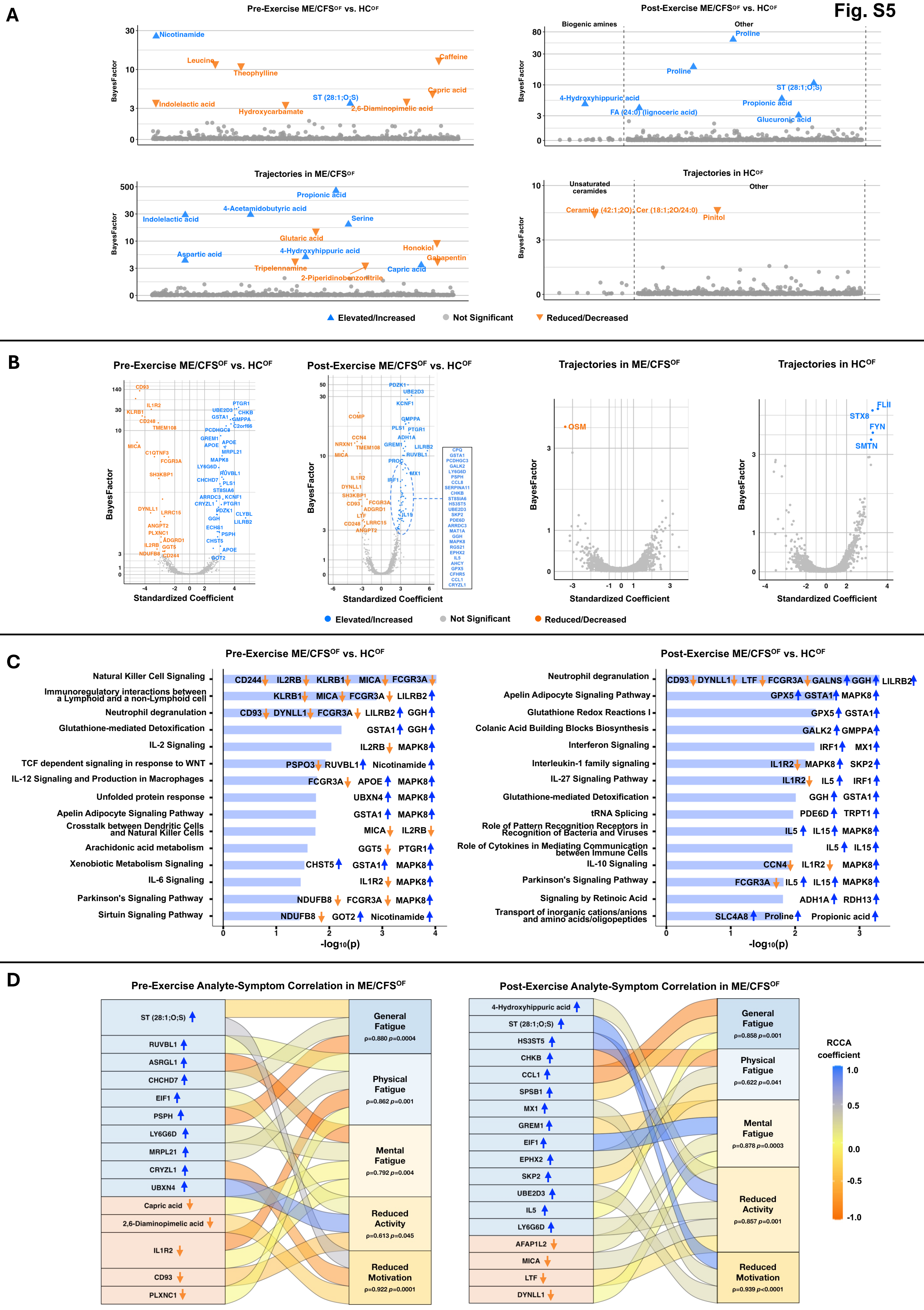

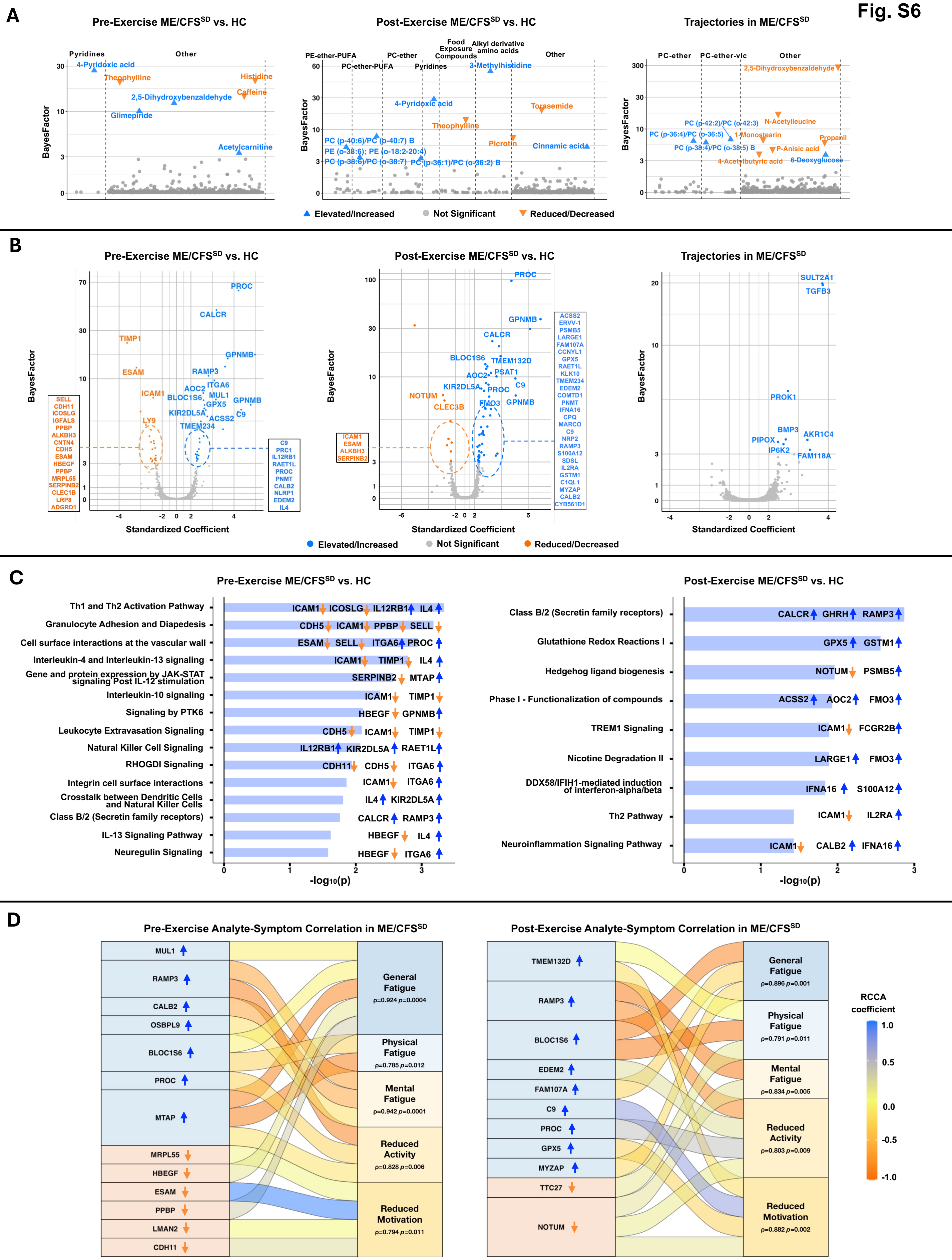

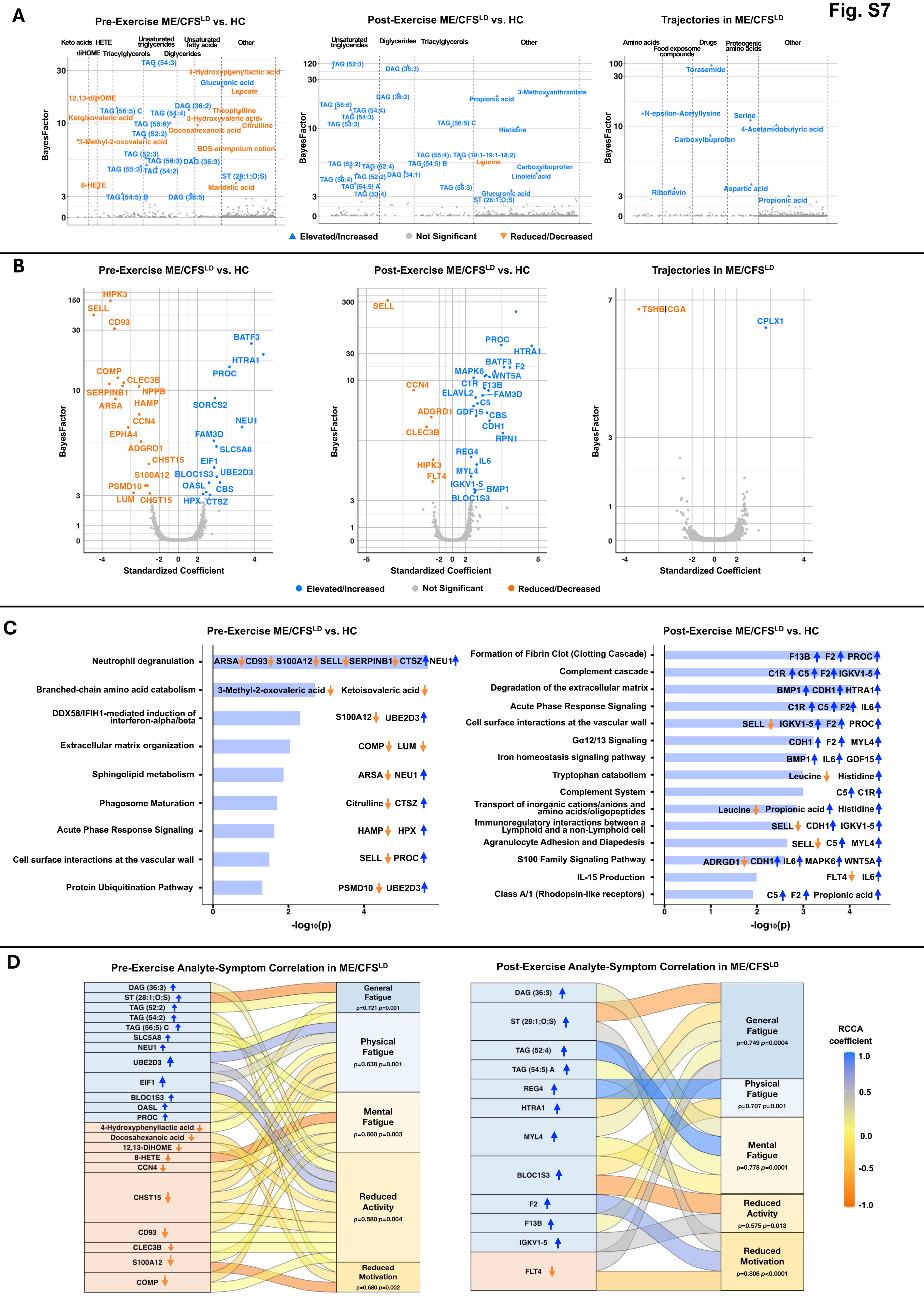
